# Differential host responses within the upper respiratory tract and peripheral blood of children and adults with SARS-CoV-2 infection

**DOI:** 10.1101/2023.07.31.23293337

**Authors:** Jillian H. Hurst, Aditya A. Mohan, Trisha Dalapati, Ian A. George, Jhoanna N. Aquino, Debra J. Lugo, Trevor S. Pfeiffer, Javier Rodriguez, Alexandre T. Rotta, Nicholas A. Turner, Thomas W. Burke, Micah T. McClain, Ricardo Henao, C. Todd DeMarco, Raul Louzao, Thomas N. Denny, Kyle M. Walsh, Zhaohui Xu, Asuncion Mejias, Octavio Ramilo, Christopher W. Woods, Matthew S. Kelly

**Affiliations:** Department of Pediatrics, Division of Infectious Diseases, Duke University School of Medicine; Durham, NC, USA; Children’s Health and Discovery Institute, Department of Pediatrics, Duke University School of Medicine; Durham, NC, USA; Medical Scientist Training Program, Department of Neurosurgery, Duke University School of Medicine; Durham, NC, USA; Medical Scientist Training Program, Department of Molecular Genetics and Microbiology, Duke University School of Medicine; Durham, NC, USA; Duke University School of Medicine; Durham, NC, USA; Children’s Clinical Research Unit, Department of Pediatrics, Duke University School of Medicine; Durham, NC, USA; Department of Pediatrics, Division of Pediatric Critical Care Medicine, Duke University School of Medicine; Durham, NC, USA; Department of Medicine, Division of Infectious Diseases, Duke University School of Medicine; Durham, NC, USA; Center for Infectious Disease Diagnostics and Innovation, Duke University School of Medicine; Durham, NC, USA; Durham Veterans Affairs Medical Center; Durham, NC, USA; Department of Biostatistics and Informatics, Duke University, Durham, NC; Duke Clinical Research Institute, Duke University School of Medicine, Durham, NC; Duke Human Vaccine Institute, Duke University School of Medicine, Durham, NC; Department of Neurosurgery, Duke School of Medicine, Durham, NC; St. Jude Children’s Research Hospital, Department of Infectious Diseases, Memphis, TN

**Author notes:** **Corresponding author:** Matthew S. Kelly, MD, MPH. These authors contributed equally to this work (Jillian H. Hurst, Aditya A. Mohan).

## Abstract

Age is among the strongest risk factors for severe outcomes from SARS-CoV-2 infection. We sought to evaluate associations between age and both mucosal and systemic host responses to SARS-CoV-2 infection. We profiled the upper respiratory tract (URT) and peripheral blood transcriptomes of 201 participants (age range of 1 week to 83 years), including 137 non-hospitalized individuals with mild SARS-CoV-2 infection and 64 uninfected individuals. Among uninfected children and adolescents, young age was associated with upregulation of innate and adaptive immune pathways within the URT, suggesting that young children are primed to mount robust mucosal immune responses to exogeneous respiratory pathogens. SARS-CoV-2 infection was associated with broad induction of innate and adaptive immune responses within the URT of children and adolescents. Peripheral blood responses among SARS-CoV-2-infected children and adolescents were dominated by interferon pathways, while upregulation of myeloid activation, inflammatory, and coagulation pathways was observed only in adults. Systemic symptoms among SARS-CoV-2-infected subjects were associated with blunted innate and adaptive immune responses in the URT and upregulation of many of these same pathways within peripheral blood. Finally, within individuals, robust URT immune responses were correlated with decreased peripheral immune activation, suggesting that effective immune responses in the URT may promote local viral control and limit systemic immune activation and symptoms. These findings demonstrate that there are differences in immune responses to SARS-CoV-2 across the lifespan, including between young children and adolescents, and suggest that these varied host responses contribute to observed differences in the clinical presentation of SARS-CoV-2 infection by age.

**One Sentence Summary:** Age is associated with distinct upper respiratory and peripheral blood transcriptional responses among children and adults with SARS-CoV-2 infection.

## INTRODUCTION

Age is one of the most important risk factors for severe illness and death from severe acute respiratory syndrome coronavirus 2 (SARS-CoV-2) infection, the etiologic agent of coronavirus disease 2019 (COVID-19). Throughout the pandemic, the majority of hospitalizations for severe COVID-19 have been among the elderly, with individuals 65 years of age or older accounting for approximately 75% of all COVID-19 deaths in the United States as of May 2023 (*1, 2*). However, the risk of severe COVID-19 increases with age even among young adults; compared to adults 18 to 29 years of age, adults 30 to 39 years of age are approximately twice as likely to be hospitalized for COVID-19 and have four times the risk of death (*3*). By comparison, children and adolescents are at relatively low risk of severe COVID-19. Between March 2020 and May 2023, the cumulative incidence of COVID-19 hospitalization among children less than 18 years of age was 198 per 100,000 individuals, while the hospitalization rate among adults 18 years of age or older was 1,734 per 100,000 individuals during this same time period (*4*). Older age is also associated with higher symptom severity and duration of illness among children, adolescents, and adults with mild COVID-19 not requiring hospitalization (*5–7*). These differences in COVID-19 outcomes by age have persisted despite the introduction of vaccines and across successive waves of infections caused by distinct SARS-CoV-2 variants (*8–10*).

Despite the established relationship between age and COVID-19 severity, an understanding of the biological or immunological factors that contribute to this association has remained elusive. Proposed hypotheses for the low risk of severe COVID-19 among children include a lower prevalence of comorbidities (*11*), more recent exposure to seasonal coronaviruses (*12*), developmental differences in expression of the receptors that mediate binding of SARS-CoV-2 to human cells (*13, 14*), and age-related differences in immunity (*15, 16*). With regard to the latter, prior studies have compared host immune responses to SARS-CoV-2 among non-hospitalized and hospitalized adults (*17, 18*), young adult and elderly patients (*19*), and adults with SARS-CoV-2 or infections caused by other respiratory viruses (*20*). Notably, we and others have observed age-associated differences in the clinical manifestations of SARS-CoV-2 infection even among children and adolescents (*7, 21, 22*), but there is a significant gap in our understanding of the extent to which immune responses to SARS-CoV-2 infection vary across the full age spectrum because of limited data from infants, children and adolescents (*20*).

To date, the majority of studies evaluating host responses to SARS-CoV-2 infection have focused on systemic responses or responses within specific immune cell subsets (*23*). The upper respiratory tract (URT) is the primary point of entry for SARS-CoV-2; thus, an improved understanding of host responses to the virus within this niche could identify biological processes that influence infection susceptibility, illness severity, and systemic immune responses to the virus. Several recent studies have described differences in host responses to SARS-CoV-2 within the nasal mucosa of children and adults (*24–26*); however, these studies were limited by their small sample sizes and combined analyses of pediatric age groups despite known differences in immune responses across infancy, childhood, and adolescence (*27*).

In this study, we investigated the URT and peripheral blood transcriptional responses of 201 individuals infected with or exposed to SARS-CoV-2 and ranging in age from 1 week to 83 years. We describe distinct gene expression profiles among uninfected children and adolescents by age, including markedly upregulated innate and adaptive immune responses within the URT associated with younger age. We further provide a detailed characterization of the URT and systemic host responses to acute SARS-CoV-2 infection among multiple pediatric age groups and adults. Within peripheral blood, we identify a shift towards innate immune cell populations with increasing age and upregulation of myeloid activation, inflammatory, and coagulation pathways only in SARS-CoV-2-infected adults. Finally, we identify associations between patient and illness characteristics and transcriptional responses within the URT and peripheral blood of SARS-CoV-2-infected subjects. This detailed analysis of host gene expression in one of the largest and most age-diverse cohorts studied to date reveals significant differences in the host response to SARS-CoV-2 infection across the lifespan. Moreover, these data provide new insights into the relationship between upper respiratory and systemic host responses to SARS-CoV-2 infection and how responses in these compartments may contribute to age-associated differences in COVID-19 symptoms and illness severity.

## Results

### Characteristics of the study population

We analyzed bulk RNA sequencing data from nasopharyngeal swab (n=132) and peripheral blood samples (n=155) collected from 160 non-hospitalized children and adolescents (<21 years of age) who were enrolled in the Biorepository of RespirAtory Virus Exposed (BRAVE) Kids study between April 1, 2020, and December 31, 2020, prior to the routine availability of COVID-19 vaccines or widespread circulation of major SARS-CoV-2 variants of concern (*7*). Median age [interquartile range (IQR)] was 11.5 (6.6, 15.7) years and 87 (54%) subjects were female (**Table 1**). Participants were classified as symptomatic SARS-CoV-2-infected (n=63, 39%; defined as presence of fever, cough, or shortness of breath for two or more days), asymptomatic SARS-CoV-2-infected (n=48, 30%), or asymptomatic SARS-CoV-2-uninfected (n=49, 31%). To investigate systemic responses to SARS-CoV-2 across the full spectrum of age, we additionally analyzed bulk RNA sequencing data from peripheral blood collected from 42 non-hospitalized adults (21 years of age or older), including 26 SARS-CoV-2-infected adults enrolled during the same time period in 2020 as the pediatric cohort. Median (IQR) age was 46.2 (40.1, 53.8) years and 21 (50%) were female (**Table 1**). These adult participants were classified as symptomatic SARS-CoV-2-infected (n=23, 55%), asymptomatic SARS-CoV-2-infected (n=3, 7%), or asymptomatic SARS-CoV-2-uninfected (n=16, 38%). The prevalence of respiratory symptoms such as cough, rhinorrhea, and nasal congestion differed among SARS-CoV-2-infected subjects by age category (p<0.0001 for all comparisons); compared to subjects 14-20 years of age (hereafter referred to as adolescents) and 21 years of age or older (adults), these symptoms were reported less frequently for children 0-5 years of age (young children) and children 6-13 years of age (school-age children). The timing of sample collection was similar across age categories relative to symptom onset among symptomatic SARS-CoV-2-infected subjects (Kruskal-Wallis test, *p*=0.14) and relative to SARS-CoV-2 diagnosis among asymptomatic SARS-CoV-2-infected subjects (Kruskal-Wallis test, *p*=0.82). Moreover, nasopharyngeal SARS-CoV-2 viral loads were similar among SARS-CoV-2-infected subjects by age group (Kruskal-Wallis test, *p*=0.37) and symptom presence (Wilcoxon rank-sum test, *p*=0.41).

**Table 1.** Characteristics of the study population.

| Subject characteristics | Young children<br>(0-5 yr, n=39) | School-age<br>children<br>(6-13 yr, n=65) | Adolescents<br>(14-20 yr, n=56) | Adults<br>(≥21 yr, n=41) | *p* |
| --- | --- | --- | --- | --- | --- |
| Median [IQR] age, years | 3.0 [1.6, 4.9] | 10.1 [8.4, 12.1] | 17.1 [15.6, 18.5] | 46.3 [39.9, 53.9] | <0.0001 |
| Female sex | 25 (64%) | 32 (49%) | 30 (54%) | 20 (49%) | 0.46 |
| Comorbidity, any | 9 (23%) | 27 (42%) | 23 (41%) | 18 (49%) | 0.12 |
| Asthma | 2 (5%) | 11 (17%) | 8 (14%) | 1 (3%) | 0.07 |
| Obesity | 8 (21%) | 18 (28%) | 18 (32%) | 6 (20%) | 0.51 |
| SARS-CoV-2 infection status |  |  |  |  | 0.08 |
| Asymptomatic SARS-CoV-2-uninfected | 10 (26%) | 22 (34%) | 17 (30%) | 15 (37%) |  |
| Asymptomatic SARS-CoV-2-infected | 14 (36%) | 20 (31%) | 14 (25%) | 3 (7%) |  |
| Symptomatic SARS-CoV-2-infected | 15 (38%) | 23 (35%) | 25 (45%) | 23 (56%) |  |
| **SARS-CoV-2 illness characteristics** |  |  |  |  |  |
| Fever | 13 (45%) | 16 (37%) | 16 (41%) | 14 (54%) | 0.59 |
| Cough | 7 (24%) | 10 (23%) | 19 (49%) | 19 (73%) | <0.0001 |
| Rhinorrhea | 4 (14%) | 1 (2%) | 8 (21%) | 14 (54%) | <0.0001 |
| Nasal congestion | 2 (7%) | 3 (7%) | 8 (21%) | 17 (65%) | <0.0001 |
| Median [IQR] nasopharyngeal viral load,<br>log<sub>10</sub> copies/mL | 5.8 [4.0, 7.1] | 5.1 [3.5, 7.0] | 5.1 [3.7, 6.8] | 5.7 [4.6, 6.8] | 0.37 |

IQR, interquartile range; mL, milliliter

### Genes expression profiles of subjects without SARS-CoV-2 infection

We first sought to describe age-associated differences in gene expression in the URT and peripheral blood of healthy children, adolescents, and adults. We thus compared the transcriptional profiles of asymptomatic SARS-CoV-2-uninfected young children (n=10), school-age children (n=22), adolescents (n=17), and adults (n=16, peripheral blood samples only). Because immune cell composition is known to vary with age (*28*), we first estimated the proportions of immune cell populations within URT and peripheral blood samples using the CIBERSORT deconvolution algorithm and the LM22 signature matrix (**Fig. 1a**) (*29*). We observed no significant differences in the proportions of immune cells in the URT of healthy children and adolescents by age. Within the peripheral blood of children, adolescents, and adults, the proportions of immune cells represented by B cells (beta regression, *p*_adj_<0.0001), CD8+ T cells (*p*_adj_=0.002), and plasma cells (*p*_adj_=0.005) decreased with increasing age, while the proportions accounted for by monocytes and macrophages (*p*_adj_<0.0001) and neutrophils (*p*_adj_=0.0009) increased. Because of these differences in immune cell populations by age, and similar differences observed in analyses by SARS-CoV-2 infection status, we adjusted all gene expression analyses for sample immune cell composition. This approach enabled us to identify genes that are differentially expressed independent of age-associated differences in immune cell proportions and that more accurately reflect activation or suppression of gene expression within these cell populations. Within the URT of SARS-CoV-2-uninfected subjects, we identified few genes that were differentially expressed between young children and school-age children and between school-age children and adolescents (**Fig. S1a**). In contrast, we identified 83 genes that were differentially expressed in young children compared to adolescents. Within peripheral blood, few genes were differentially expressed across pediatric age groups or in comparisons of pediatric age groups to adults after accounting for differences in immune cell proportions (**Fig. S1b**).

**Fig. 1.**
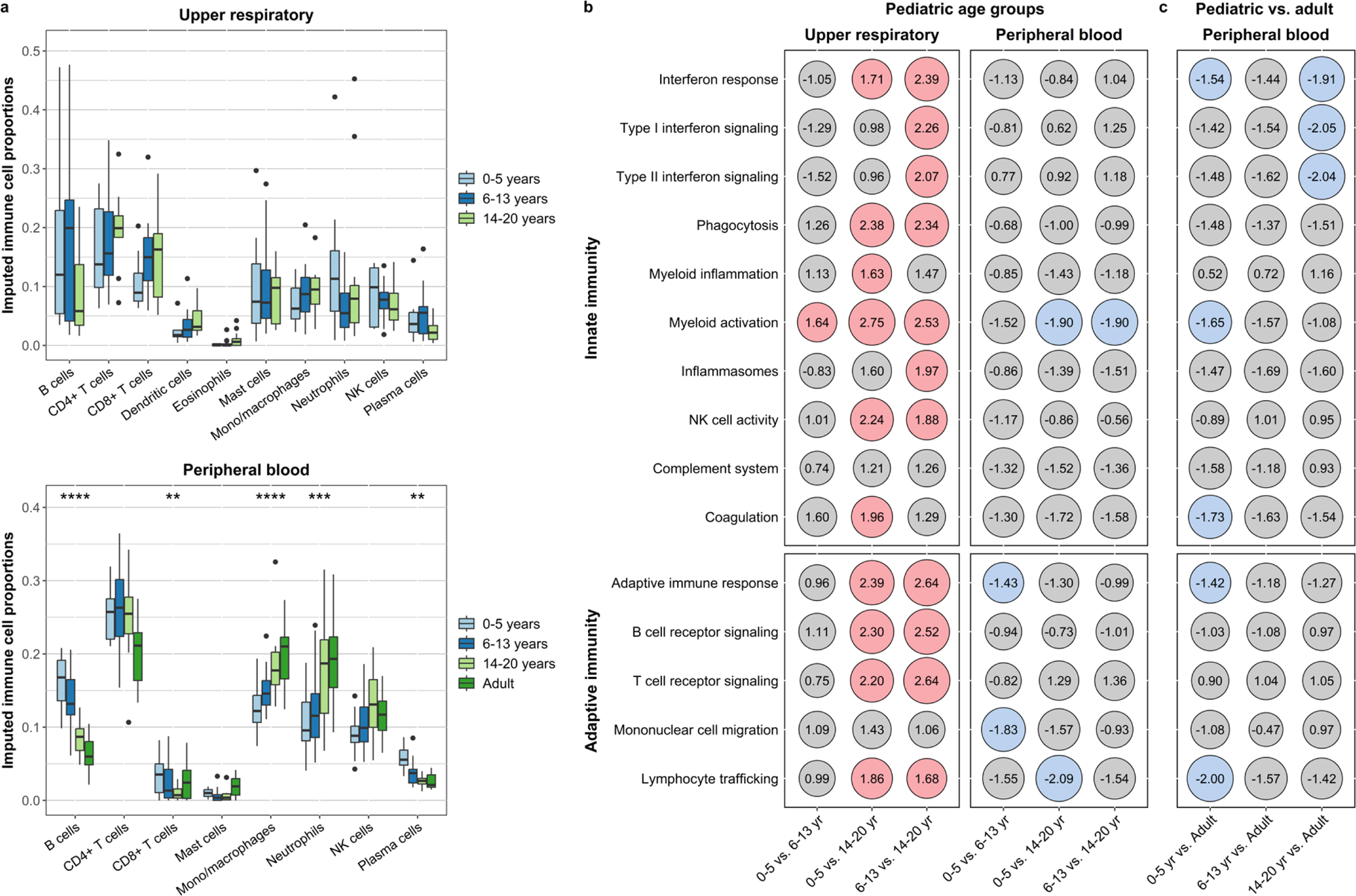
Transcriptional profiles within the upper respiratory tract and peripheral blood of SARS-CoV-2-uninfected children, adolescents, and adults. Bulk RNA sequencing was used to compare the transcriptional profiles of healthy young children (0-5 years), school-age children (6-13 years), adolescents (14-20 years), and adults (≥21 years, peripheral blood samples only). **a.** Box and whisker plots depict proportions of immune cell populations in the upper respiratory tract and peripheral blood of SARS-CoV-2-uninfected children and adolescents, imputed using CIBERSORT. Lines splitting the boxes correspond to median values while box edges represent the 25^th^ and 75^th^ percentiles with outliers shown as single points. Proportions of immune cell populations were compared by age (modeled as a continuous variable) using beta regression, with all analyses corrected for multiple comparisons (*, *p*_adj_<0.05; **, *p*_adj_<0.01; ***, *p*_adj_<0.001; ****, *p*_adj_<0.0001). Only immune cell populations identified in at least 25% of samples are shown. **b.** and **c.** Differential expression of gene modules in upper respiratory and peripheral blood samples from pediatric age groups (**b.**) and in peripheral blood samples from children and adolescents compared to adults (**c.**). Below each column, the age group listed first represents the group of interest while the age group listed second is the reference group. Numbers within each circle indicate the normalized enrichment score (NES) calculated through gene set enrichment analysis. Colored circles indicate differential expression (*p*_adj_<0.05) of immune modules across the comparison groups (red, upregulation in the age group of interest; blue, downregulation in the age group of interest). Gene set enrichment analyses were adjusted for sex, sequencing batch, and imputed sample immune cell proportions. (NK, natural killer)

To compare expression of biological pathways by age, we used the gene sets defined by the NanoString nCounter^®^ Host Response Panel to identify groups of co-expressed genes that share a similar function (hereafter referred to as modules). We then used fast gene set enrichment analysis (FGSEA) to evaluate for differential expression of these modules across age groups (*30, 31*). We observed few differences between young children and school-age children; however, compared to adolescents, young children and school-age children exhibited higher expression of multiple innate immune modules (e.g., interferon signaling, phagocytosis, myeloid cell activation and inflammation, and NK cell activity) and adaptive immune modules (e.g., B cell receptor and T cell receptor signaling, and lymphocyte trafficking) within the URT (**Fig. 1b, Table S1**). Peripheral blood expression of many of these same modules was similar across pediatric age groups; however, lower expression of genes associated with myeloid cell activation was observed with decreasing age among children and adolescents (**Fig. 1b, Table S2**). Compared to adults, pediatric age groups tended to have lower peripheral blood expression of interferon signaling pathways and genes associated with coagulation and myeloid cell and inflammasome activation (**Fig. 1c, Table S2**). Taken together, these data indicate that younger age is associated with greater baseline activation of immune cell populations within the URT which may prime children for more rapid or robust immune responses to exogenous respiratory pathogens.

### Gene expression profiles in SARS-CoV-2-infected children, adolescents, and adults

We next compared host transcriptional responses in SARS-CoV-2-infected subjects and healthy subjects without SARS-CoV-2. We first used CIBERSORT to compare the proportions of immune cell populations in URT and peripheral blood samples by SARS-CoV-2 infection status. SARS-CoV-2-infected subjects had a lower proportion of dendritic cells (*p*_adj_=0.02) and a higher proportion of monocytes and macrophages (*p*_adj_=0.03) within the URT (**Fig. S2a**) and higher proportion of plasma cells (*p*_adj_=0.03) in peripheral blood (**Fig. S2b**). Among pediatric subjects, we found that acute SARS-CoV-2 infection triggered a robust transcriptional response in the URT characterized by upregulation of gene expression for complement pathway components (e.g., *C1QB*), macrophage activity (e.g., *CD163, MSR1*), interferon-inducible genes (e.g., *IFI6, IFI44L, NRIR, SIGLEC-1*), immune cell trafficking (e.g., *CCL2, CXCL11, CXCL10*)), and genes related to cytotoxic immune responses (e.g., *GZMB, LAG3*) (**Fig. 2a**). Similarly, gene set enrichment analyses demonstrated upregulation of a broad array of innate and adaptive immune modules within the URT of SARS-CoV-2-infected subjects (**Fig. 2a, Table S3**). Similar transcriptional responses were observed in the peripheral blood of SARS-CoV-2-infected subjects; however, expression of innate and adaptive immune modules was generally upregulated to a lesser degree in peripheral blood than in the URT. Further, we observed upregulation of Nod-like receptor and RNA and DNA sensing pathways exclusively in peripheral blood, while chemokine signaling, NFκB signaling, and T cell receptor signaling were only upregulated in the URT (**Fig. 2b**, **Table S3**).

**Fig. 2.**
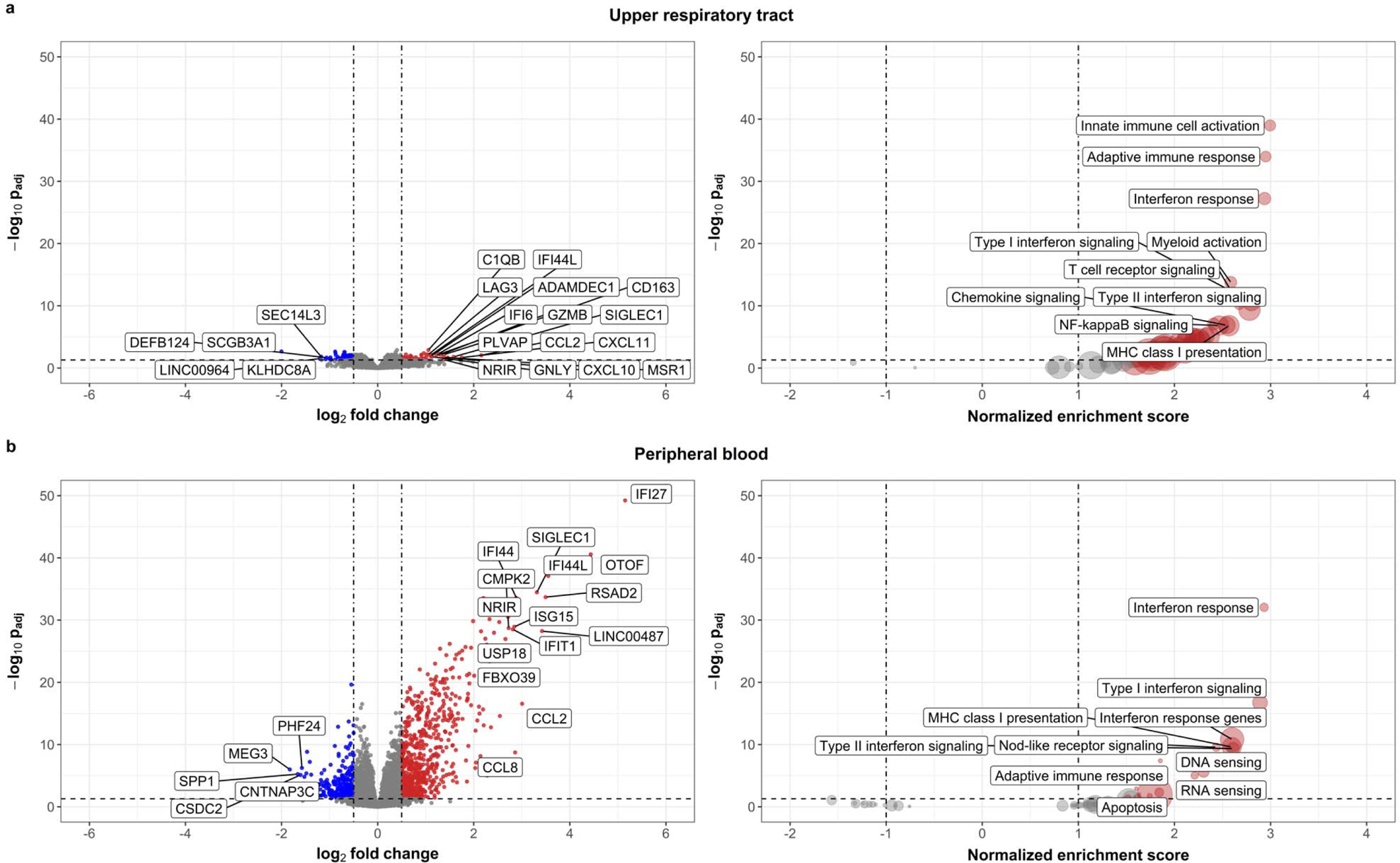
Differential host gene expression in upper respiratory and peripheral blood samples associated with SARS-CoV-2 infection among children, adolescents, and adults. Bulk RNA sequencing was used to compare the upper respiratory and peripheral blood transcriptional profiles of young children (0-5 years), school-age children (6-13 years), adolescents (14-20 years), and adults (≥21 years, peripheral blood only) by SARS-CoV-2 infection status. **a.** Volcano plot depicting differential expression of genes in the upper respiratory tracts of SARS-CoV-2-infected compared to uninfected children and adolescents (*p*_adj_<0.05). The 15 most differentially upregulated and 5 most downregulated genes in SARS-CoV-2-infected subjects based on log2-fold change are labeled. **b.** Volcano plot depicting differential expression of gene modules in upper respiratory samples by SARS-CoV-2 infection status. The 10 modules with the highest normalized enrichment scores (NES; upregulated in SARS-CoV-2 infection) are labeled; no modules were downregulated in SARS-CoV-2-infected subjects. **c.** Volcano plot depicting differential expression of genes in the peripheral blood of SARS-CoV-2-infected compared to uninfected children, adolescents, and adults. The 15 most differentially upregulated and 5 most downregulated genes in SARS-CoV-2-infected subjects based on log2-fold change are labeled. **d.** Volcano plot depicting differential expression of gene modules in peripheral blood by SARS-CoV-2 infection status. The 10 modules with the highest NES (upregulated in SARS-CoV-2-infected subjects) are labeled; no modules were downregulated in SARS-CoV-2-infected subjects. All analyses shown were adjusted for sex, sequencing batch, age group, and imputed sample immune cell proportions. (MHC, major histocompatibility complex)

### Differences in upper respiratory transcriptional responses to SARS-CoV-2 infection by age

In order to assess the extent to which immune responses to SARS-CoV-2 within the URT differ based on age, we compared transcriptional responses between infected and uninfected participants by age group. Using CIBERSORT, we found that there were no differences in the proportions of immune cell populations within URT samples by age (**Fig. 3a**). We then characterized changes in URT gene and immune module expression associated with SARS-CoV-2 infection within each age group by comparing transcriptional profiles of SARS-CoV-2-infected subjects to those of uninfected subjects within that same age group. Gene set enrichment analyses identified upregulation of both innate and adaptive immune modules in the URT across all pediatric age groups, including interferon responses, myeloid cell activation, inflammasome signaling, and B and T cell receptor signaling (**Fig. 3b, Table S4**). Moreover, a number of these modules, including those corresponding to interferon signaling, myeloid inflammation, and adaptive immune cell trafficking, were upregulated to a greater degree in young children than in school-age children or adolescents. We also observed a larger number of differentially expressed genes in SARS-CoV-2-infected versus uninfected young children (57 genes) relative to other pediatric age groups (school-age children, 5 genes; adolescents, 10 genes), with differential expression of chemokines (e.g., *CCL2*, *CXCL10*, *CXCL11*) and genes involved in interferon signaling (e.g., *IFITM3*, *IFI6*) and antigen presentation (e.g., *HLA-DPA1*) (**Fig. S3**). Notably, adolescents were the only pediatric age group in which upregulation of complement signaling (e.g., *C1QB*, *C1QC*) was observed in the setting of SARS-CoV-2 infection. Taken together, these findings suggest that younger age is associated not only with greater baseline activation of innate and adaptive immune pathways within the URT but also with more robust upregulation of these pathways in response to SARS-CoV-2 infection.

**Fig. 3.**
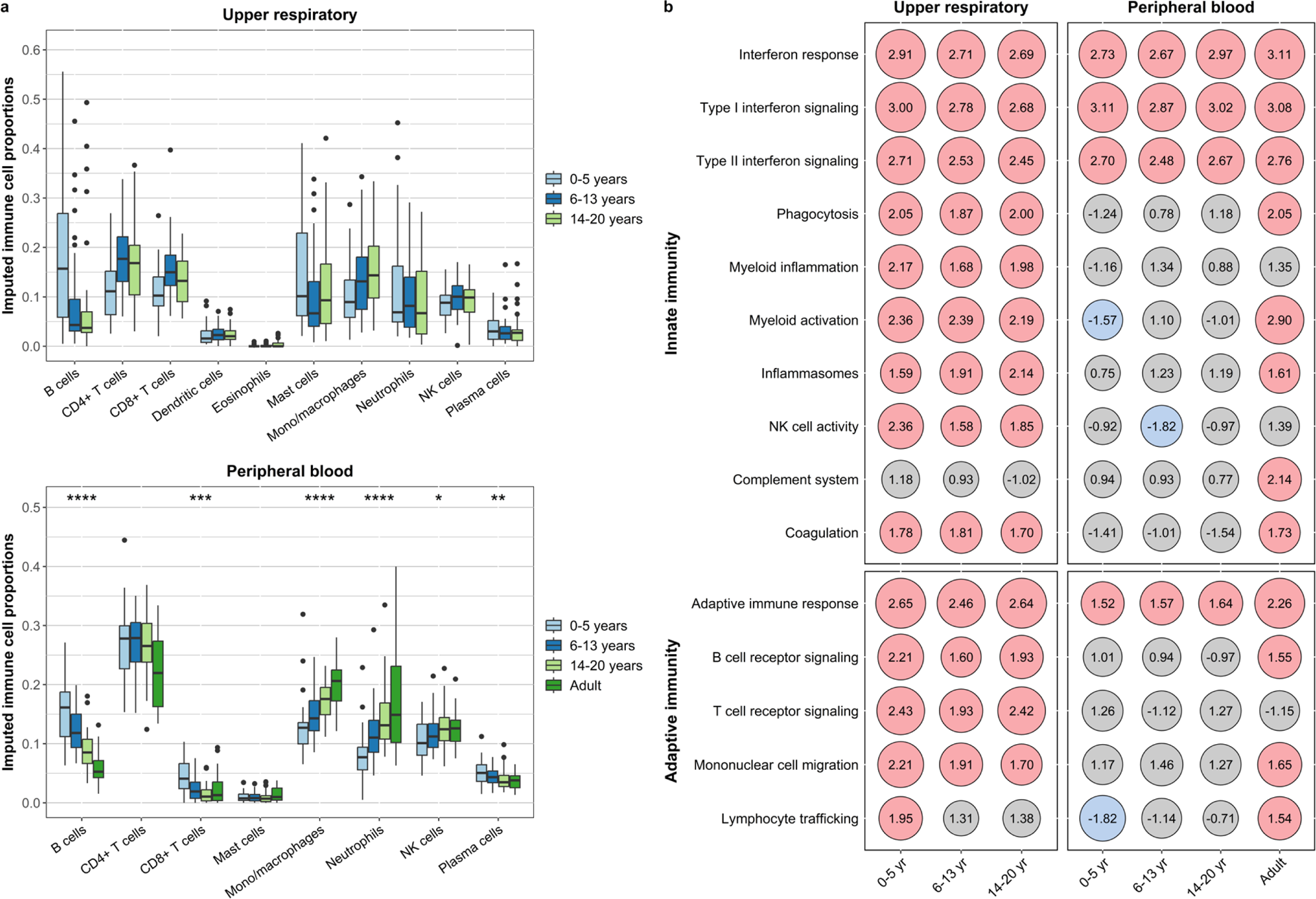
Transcriptional profiles associated with SARS-CoV-2 infection by age group. Bulk RNA sequencing was used to compare the upper respiratory and peripheral blood transcriptional profiles of young children (0-5 years) school-age children (6-13 years), adolescents (14-20 years), and adults (≥21 years; peripheral blood only). **a.** Box and whisker plots depict proportions of immune cell populations in upper respiratory samples and peripheral blood of SARS-CoV-2-infected subjects, imputed using CIBERSORT. Lines splitting the boxes correspond to median values while box edges represent the 25^th^ and 75^th^ percentiles with outliers shown as single points. Proportions of immune cell populations were compared by age (modeled as a continuous variable) using beta regression, with all analyses corrected for multiple comparisons (*, *p*_adj_<0.05; **, *p*_adj_<0.01; ***, *p*_adj_<0.001; ****, *p*_adj_<0.0001). Only immune cell populations identified in at least 25% of samples are shown. **b.** Differential expression of gene modules in the upper respiratory tract and peripheral blood associated with SARS-CoV-2 infection by age group. Within each age group listed below the columns, module expression among SARS-CoV-2-infected subjects was compared to that of uninfected subjects in that same age group. Numbers within each circle indicate the normalized enrichment score (NES) calculated through gene set enrichment analysis. Colored circles indicate differential expression (*p*_adj_<0.05) of gene modules by SARS-CoV-2 infection status (red, upregulated in SARS-CoV-2-infected subjects; blue, downregulated in SARS-CoV-2-infected subjects). Gene set enrichment analyses were adjusted for sex, sequencing batch, and imputed sample immune cell proportions. (NK, natural killer)

### Differences in peripheral blood transcriptional responses to SARS-CoV-2 infection by age

To identify age-associated differences in systemic host responses to SARS-CoV-2 infection, we compared transcriptional profiles in peripheral blood from SARS-CoV-2-infected and uninfected subjects by age group. Using CIBERSORT to infer immune cell proportions, we identified age-associated differences in multiple immune cell populations in the peripheral blood of infected subjects (**Fig. 3a**). In particular, the proportions of immune cells represented by specific innate cell populations increased with age (monocytes and macrophages, *p*_adj_<0.0001; NK cells, *p*_adj_=0.02; neutrophils, *p*_adj_<0.0001), while the proportions of adaptive immune cell populations decreased with age (B cells, *p*_adj_<0.0001; CD8^+^ T cells, *p*_adj_=0.0003; plasma cells, *p*_adj_=0.001). Notably, the age-associated differences in immune cell populations among SARS-CoV-2-infected subjects were similar to those observed in healthy subjects. Gene set enrichment analysis of data from peripheral blood samples revealed increases in interferon signaling across all age groups of SARS-CoV-2-infected subjects, though adults tended to exhibit greater upregulation of these modules than pediatric age groups (**Fig. 3b, Table S5**); this pattern was also reflected in expression of individual genes involved in interferon responses (**Fig. S4**). Adults also demonstrated greater upregulation of pathways associated with innate immune responses compared to children and adolescents (**Fig. 3b, Table S5**). Most notably, upregulation of modules corresponding to myeloid cell activation, inflammasomes, NK cell activity, the complement system, and coagulation was observed exclusively in adults, with many of these same pathways being downregulated in SARS-CoV-2-infected children and adolescents. Finally, although the transcriptional activity of adaptive immune cells was generally higher in the peripheral blood of SARS-CoV-2-infected adults relative to pediatric age groups, adults had lower proportions of B cells and plasma cells in peripheral blood. These findings are consistent with a more robust systemic immune response to SARS-CoV-2 infection in adults relative to children and adolescents that includes upregulation of pathways associated with innate immunity, inflammation, and coagulation.

### Differences in transcriptional responses to SARS-CoV-2 infection by illness characteristics

Because we and others have previously observed marked differences in the symptoms of SARS-CoV-2 infection across age groups (*7, 32–34*), we next evaluated associations between illness characteristics and the URT and peripheral blood transcriptional responses of SARS-CoV-2-infected subjects (**Fig. 4, Tables S6 and S7**). We found that a high nasopharyngeal viral load (≥10^6^ copies/mL) was associated with broad upregulation of innate and adaptive immune modules within the URT, with particularly high expression of modules corresponding to interferon signaling, Nod-like receptor signaling, and adaptive immune responses, as well as upregulation of several innate and adaptive immune modules in peripheral blood. We also observed upregulation of many innate and adaptive immune modules in URT samples from subjects with asthma, whereas obesity was associated with downregulation of several immune modules in the URT, particularly those corresponding to interferon signaling and innate immune responses. Fever and cough were associated with upregulation of interferon signaling and other innate and adaptive immune modules in peripheral blood; however, unexpectedly, fever was associated with broad downregulation of innate and adaptive immune responses within the URT, with the largest decreases in gene expression observed for interferon signaling, innate immune cell activation, and myeloid inflammation. Upper respiratory symptoms (i.e., rhinorrhea and nasal congestion) were associated with downregulation of both innate and adaptive immune responses in the URT, with few differences noted in peripheral blood immune module expression. Taken together, these data suggest that symptoms localizing outside of the URT, such as fever and cough, are associated with less robust immune responses to SARS-CoV-2 in the URT and more robust systemic immune responses. In contrast, upper respiratory symptoms are associated with blunted innate and adaptive immune responses to SARS-CoV-2 in the URT without associated systemic immune activation.

**Fig. 4.**
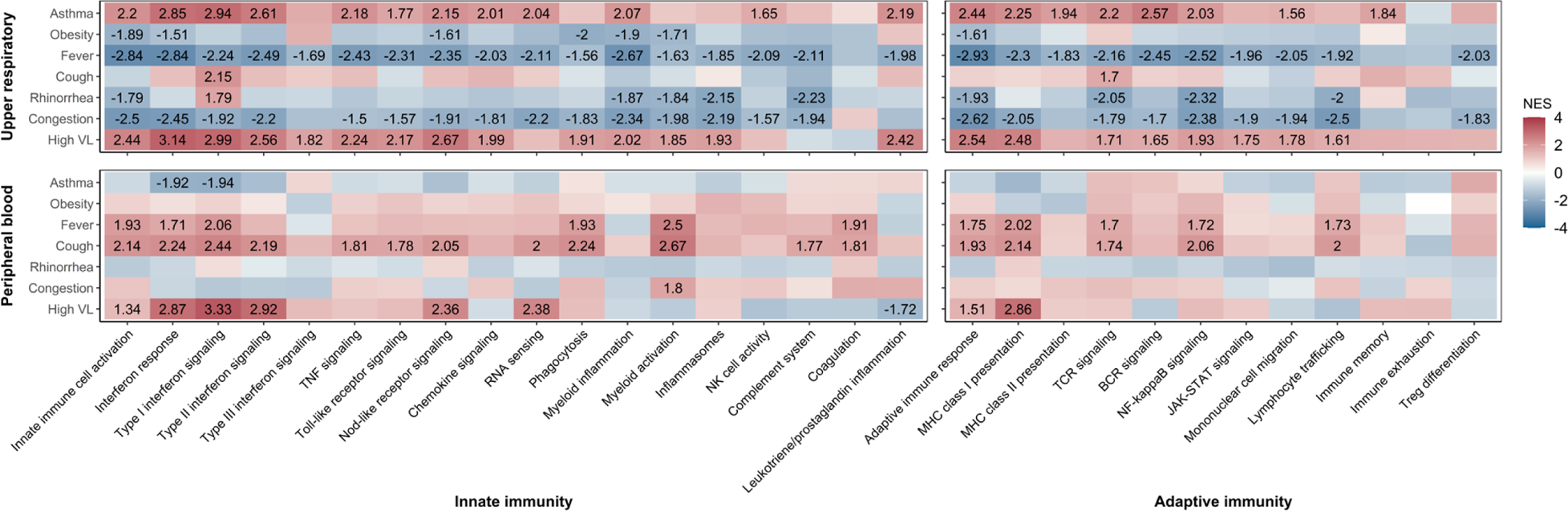
Differential expression of immune modules in SARS-CoV-2 infection by subject and illness characteristics. Heatmaps depict results of gene set enrichment analysis of upper respiratory (children and adolescents) and peripheral blood (children, adolescents, and adults) bulk RNA sequencing data for comparisons of SARS-CoV-2-infected subjects with and without specific comorbidities (asthma, obesity); symptoms present during acute infection (fever, cough, rhinorrhea, congestion); and with high versus low nasopharyngeal viral loads (high viral load defined as ≥10^6^ copies/mL). Each column corresponds to a gene module comprised of co-expressed genes that share a similar function. The numbers within blocks represent normalized enrichment scores (NES) for comparisons of gene module expression that were statistically significant (*p*_adj_<0.05). The color of each block indicates the direction of the change in expression (red, upregulated in the condition of interest; blue, downregulated in the condition of interest). Analyses were adjusted for sex, sequencing batch, age group, and imputed sample immune cell proportions.

### Correlations between host response within the upper respiratory tract and peripheral blood

Given that the URT serves as the primary entry point for SARS-CoV-2 among exposed individuals, we hypothesized that a strong antiviral response within the URT would be associated with blunted systemic immune responses. We therefore evaluated correlations between expression of immune modules in paired URT and peripheral blood samples collected from children and adolescents. We observed strong positive correlations between expression of both interferon signaling and RNA sensing pathways in URT and peripheral blood samples (**Fig. 5, Table S8**). In contrast, higher URT expression of other innate immune modules, including those corresponding to phagocytosis, myeloid activation, and NK cell activity, was associated with lower expression of similar innate immune pathways within peripheral blood. Similarly, URT expression of adaptive immune modules, including B cell receptor signaling, JAK-STAT signaling, NK cell trafficking, and immune memory pathways, was negatively correlated with peripheral blood expression of several innate immune modules, including those for myeloid cell activity, lymphocyte trafficking, and inflammatory modules corresponding to Nod-like receptor signaling and inflammasome activity. Finally, to evaluate the extent to which age influences these correlations between immune responses in the URT and peripheral blood, we performed similar analyses for each pediatric age group (**Fig. S5, Table S8**). Interferon signaling in the URT and peripheral blood were positively correlated in all age groups. However, fewer correlations between immune responses in the URT and peripheral blood were observed among young children and adolescents than among school-age children. Interestingly, school-age children were noted to have a lower prevalence of systemic symptoms both in the original description of the BRAVE Kids cohort and in the subset of subjects analyzed herein (**Table 1**) (*7*). These findings suggest that there are age-associated differences in the coordination of local and systemic immune responses to SARS-CoV-2 that contribute to the varied clinical manifestations of this infection among pediatric age groups.

**Fig. 5.**
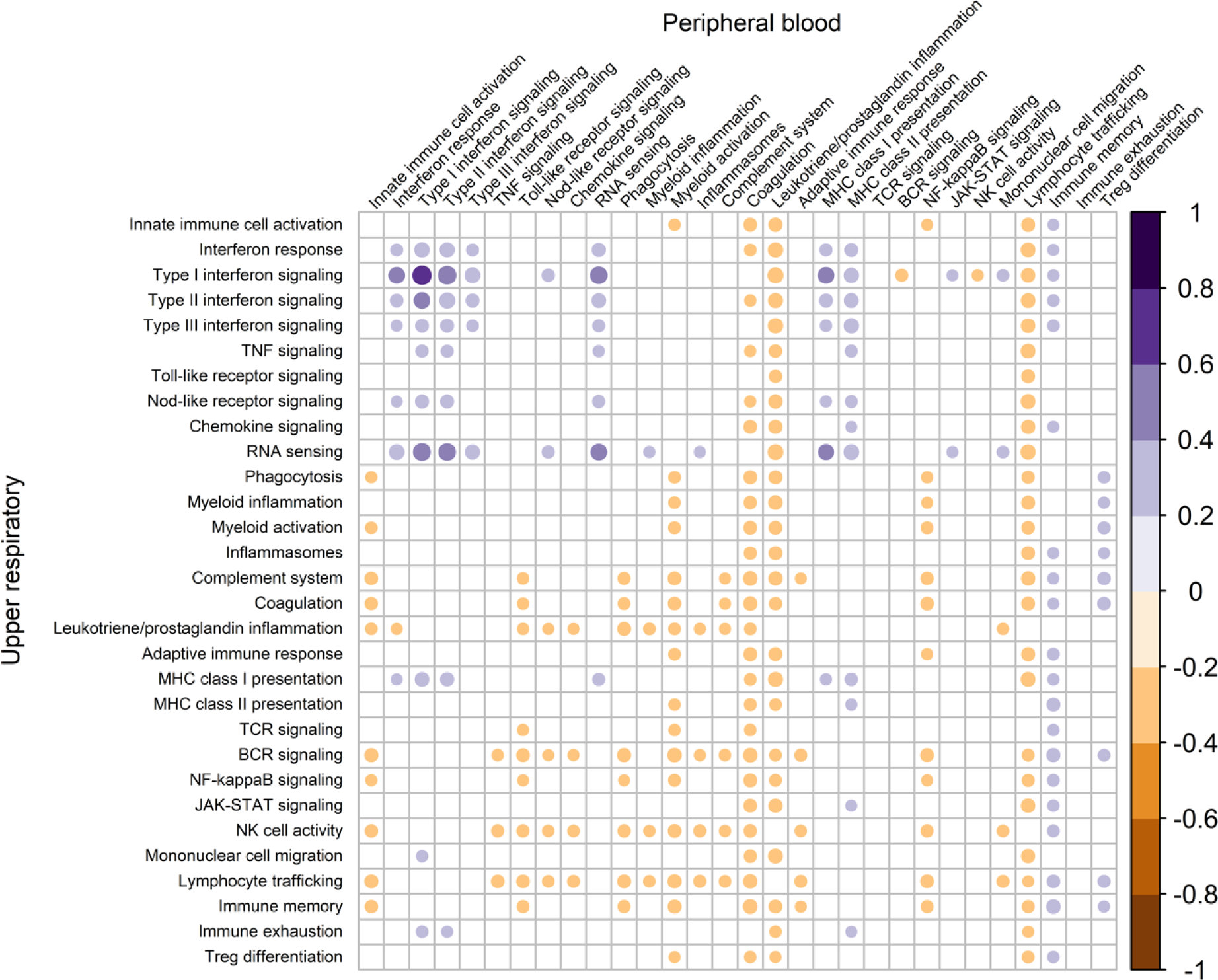
Correlations between immune module expression in the upper respiratory tract and peripheral blood of SARS-CoV-2-infected children and adolescents. Single-sample gene set enrichment analysis was used to calculate enrichment scores for gene modules in paired upper respiratory and peripheral blood samples from SARS-CoV-2-infected children and adolescents. Pearson’s correlation coefficients were then calculated to evaluate for linear relationships between expression of these modules in the upper respiratory tract and peripheral blood of the same individual. Positive correlations are displayed in purple and negative correlations are displayed in orange. The size and color of each circle corresponds to the strength of the correlation; only statistically significant correlations (*p*<0.05) are shown.

## DISCUSSION

In this study, we performed bulk RNA sequencing of URT and peripheral blood samples from 201 individuals across the lifespan, evaluating host gene expression in one of the largest and most age-diverse cohorts studied to date. Further, unlike many prior studies, our cohort only included patients who did not require hospitalization for COVID-19 and is therefore representative of the vast majority of SARS-CoV-2 infections, particularly among children and adolescents. We demonstrate that there are distinct mucosal and systemic host responses to SARS-CoV-2 infection across the age spectrum. Moreover, we describe relationships between compartment-specific immune responses in the URT and peripheral blood that likely contribute to observed differences in the clinical presentation of SARS-CoV-2 infection among children, adolescents, and adults.

One of the most striking epidemiological features of the COVID-19 pandemic has been the variable disease severity among different age groups. In particular, children and adolescents have been at low risk of severe disease, suggesting that important insights can be gained by studying the immune responses of pediatric age groups to SARS-CoV-2 infection. Prior studies of systemic host responses to SARS-CoV-2 infection have noted striking differences by age, but these studies have included relatively few children and adolescents, and most have not evaluated for differences between pediatric age groups. Using single-cell RNA sequencing, Yoshida and colleagues reported that 19 children and adolescents (<18 years of age) exhibited a more naïve and less cytotoxic T cell response to SARS-CoV-2 infection in peripheral blood compared to 18 adults (*24*). In a longitudinal household transmission study, Vono and colleagues observed that children and adolescents (n=16, age range: 0.8 to 16 years) mounted a faster, more transient systemic immune response to SARS-CoV-2 infection than 21 adults, though children and adults exhibited similar expression of antiviral immune response genes, including genes associated with interferon responses, immune cell activation, and inflammation (*35*). Finally, Pierce and colleagues reported that children, adolescents, and young adults (<24 years of age) hospitalized for COVID-19 (n=65) had lower serum concentrations of inflammatory cytokines and muted peripheral blood cellular immune responses compared to hospitalized adults 24 years of age or older (n=60) (*36*). In one of the largest analyses of SARS-CoV-2-infected children and adolescents conducted to date, we observe that pediatric age groups have less systemic activation of several innate immune and inflammatory pathways compared to adults. Notably, we found that SARS-CoV-2 infection was associated with upregulation of genes corresponding to myeloid cell activation in the peripheral blood of adults, while expression of these genes was generally suppressed among SARS-CoV-2-infected children and adolescents. Prior studies suggest that myeloid cell dysregulation plays an important role in the pathogenesis of severe COVID-19 (*37–39*); thus, the differing expression of myeloid cell-associated genes in response to SARS-CoV-2 infection that we observed among children and adults could contribute to the varied illness presentation in these age groups. Moreover, consistent with the clinical observation that children and adolescents with COVID-19 are at lower risk of thrombotic complications than adults (*40*), we show that even mild SARS-CoV-2 infection is associated with upregulation of genes associated with coagulation among adults, while these pathways are downregulated in SARS-CoV-2 infection occurring among children and adolescents.

The URT is the primary entry point for SARS-CoV-2 and most other respiratory viruses; thus, mucosal immunity within this ecological niche represents the first opportunity for viral control (*41*). Analyzing upper respiratory samples from healthy children and adolescents without SARS-CoV-2 infection, we found that children 13 years of age or younger had higher expression of a broad range of both innate and adaptive immune pathways compared to adolescents. This observation is consistent with findings from the few prior studies that investigated age-associated differences in mucosal immune responses in healthy individuals. Yoshida and colleagues observed that URT epithelial cells from healthy children were in an interferon-activated state relative to cells from healthy adults (*24*). Similarly, Loske and colleagues performed single-cell RNA sequencing of nasopharyngeal samples from healthy children (n=18) and adults (n=23) and found increased expression of pattern recognition receptors and higher abundances of macrophages and dendritic cells in airway epithelial cells from children relative to adults (*42*). Taken together, our findings and those of these studies suggest that younger age is associated with higher baseline URT immune tone that may enable more rapid or robust responses to exogenous respiratory pathogens.

Prior studies suggest that children generate a more vigorous mucosal immune response to SARS-CoV-2 infection than adults. Pierce and colleagues performed bulk RNA sequencing of nasopharyngeal samples from 12 children and 27 adults with SARS-CoV-2 infection and observed higher expression of interferon signaling, inflammasome, and innate immune pathways among children relative to adults (*25*). Additionally, in the aforementioned study by Loske and colleagues, children with SARS-CoV-2 infection had higher URT levels of interferons and chemokines than infected adults (*42*). Findings from animal models also suggest that age influences the strength and characteristics of mucosal immune responses to SARS-CoV-2. Oishi and colleagues found that aged hamsters generated less robust pulmonary innate immune and repair responses to SARS-CoV-2 infection than younger animals, while older age was associated with diminished adaptive immune responses and increased expression of inflammatory pathways (*43*). Similarly, using a ferret model that included animals of different ages, Kim and colleagues found the oldest animals had higher expression of genes associated with immune cell activation and inflammatory responses, while younger animals had greater expression of tissue remodeling and repair pathways (*44*). In the current study, we found that children less than 5 years of age generally mounted more robust URT interferon and innate and adaptive immune responses to SARS-CoV-2 infection than older children or adolescents, indicating that age-associated differences in the mucosal immune response to SARS-CoV-2 infection exist even within the pediatric population.

An effective mucosal immune response to respiratory viruses has the potential to limit viral replication and reduce systemic immune activation, with the URT serving as a physical barrier to infection and as the first point of contact with the immune system (*42, 45*). Recent results from preclinical and early phase clinical studies of intranasal vaccines for SARS-CoV-2 and other respiratory pathogens demonstrate that mucosal pathogen exposures have a profound impact on systemic immune responses (*46–48*). Further, several prior studies demonstrated a relationship between mucosal immune responses and the severity of respiratory viral infections. In an influenza household transmission study, Oshansky and colleagues found associations between age, nasal cytokine response, and disease severity, with innate immune responses in the respiratory mucosa being the strongest predictor of clinical outcomes (*49*). In a study of 37 children with rhinovirus or respiratory syncytial virus (RSV) lower respiratory tract infection, García and colleagues found that nasal cytokine concentrations were inversely correlated with disease severity, suggesting a protective effective of a robust immune response in the URT (*50*). Similarly, Taveras and colleagues observed that children with mild RSV infection had higher levels of mucosal interferons compared to children with more severe disease (*51*). Smith and colleagues measured viral load and cytokine levels in paired nasopharyngeal and plasma samples from 49 adults hospitalized for COVID-19. They found that higher disease severity was associated with lower URT interferon expression and higher plasma levels of inflammatory cytokines, suggesting that a comparatively muted URT immune response may result both in a heightened systemic immune response and worse clinical outcomes (*52*). Though our cohort consisted of non-hospitalized individuals with mild SARS-CoV-2 infection, we found that the presence of constitutional symptoms, such as fever, was associated with diminished innate and adaptive responses in the URT and increased systemic immune activation. Moreover, among children and adolescents with paired URT and peripheral blood samples, greater upregulation of innate and adaptive immune modules in the URT was associated with less systemic activation of innate immune and inflammatory pathways. These findings suggest that strong immune responses to SARS-CoV-2 in the URT may promote local control of the virus and limit development of systemic immune responses and associated constitutional symptoms. Finally, our findings suggest that coordination of mucosal and systemic immune responses differs among pediatric age groups and has the potential to influence the clinical presentation of SARS-CoV-2 infection. Future studies evaluating paired URT and blood samples from patients with varied COVID-19 severity and with other respiratory infections will be needed to further delineate relationships between immune responses in these two compartments.

Our study has several strengths and weaknesses. Strengths of the study include the large sample size and inclusion of individuals across a wide age spectrum. Additionally, this study focused on an outpatient population with mild SARS-CoV-2 infection that was enrolled prior to the routine availability of COVID-19 vaccines and the emergence of major SARS-CoV-2 variants. Thus, this study provides an opportunity to compare mucosal and systemic host responses to the virus across age groups within a population that was largely naïve to SARS-CoV-2 at the time of study recruitment. Weaknesses of the study include a lack of severe COVID-19 cases, the inclusion of which could have provided insights into host responses associated with poor outcomes among different age groups. Additionally, our study relied on bulk RNA sequencing of the URT and peripheral blood samples, which supported compartment-specific comparisons but precluded our ability to evaluate gene expression within specific tissues or cell types and immune cell subsets. Finally, analyses of URT gene expression were limited to children and adolescents because adult nasopharyngeal samples were collected in a medium that did not sufficiently preserve RNA.

In conclusion, we found that age was strongly associated with mucosal and systemic host responses to mild SARS-CoV-2 infection. Among children and adolescents, younger age was associated with a greater degree of URT immune activation in uninfected children and with a more robust URT immune activation in response to SARS-CoV-2 infection. Correspondingly, children and adolescents exhibited a relatively muted systemic immune response to SARS-CoV-2 infection compared to adults. Finally, we identified mucosal and systemic host responses associated with specific COVID-19 symptoms and provide evidence that strong mucosal immune responses are associated with less systemic immune activation. Our findings demonstrate that host responses to SARS-CoV-2 vary substantially by age, including among children and adolescents, and contribute to observed differences in the clinical presentation of COVID-19 across the lifespan.

## MATERIALS AND METHODS

### Study Design

The children and adolescents (<21 years) included in these analyses were enrolled in the Biospecimens from RespirAtory Virus-Exposed Kids (BRAVE Kids) study, a prospective cohort study of individuals with confirmed SARS-CoV-2 infection or close contact with an individual with confirmed infection (*7*). Participants were identified either through presentation to the health system or through identification of a close contact with PCR-confirmed SARS-CoV-2 infection. We administered symptom questionnaires by telephone at enrollment and at 7, 14, and 28 days after enrollment or until subjects reported resolution of all symptoms. Nasopharyngeal samples were collected with nylon flocked swabs (Copan Italia, Brescia, Italy) and placed into RNAProtect (Qiagen, Hilden, Germany). Whole blood samples were collected into PAXgene blood RNA tubes (Qiagen). The adults (≥21 years) included in these analyses participated in the Molecular and Epidemiological Study of Suspected Infection (MESSI) study. Nasopharyngeal samples were collected using prepackaged kits containing nylon flocked swabs and viral transport medium (Dasky Life Science, Ningbo, China). Whole blood samples were collected into PAXgene blood RNA tubes (Qiagen). Detailed symptom data were similarly collected from MESSI study participants using serial questionnaires. Participants from both studies were classified as SARS-CoV-2-infected if the virus was detected by PCR testing of nasopharyngeal swabs collected for clinical or research purposes. All samples from SARS-CoV-2-infected subjects included in the analyses presented herein were collected within 14 days of symptom onset (symptomatic SARS-CoV-2 infection) or diagnosis (asymptomatic SARS-CoV-2 infection). Additionally, analyses were limited to individuals with SARS-CoV-2 infection who did not require hospitalization, had no prior history of COVID-19 vaccination, and who were enrolled between April 1, 2020, and December 31, 2020, prior to widespread circulation of major SARS-CoV-2 variants of concern. Subjects classified as SARS-CoV-2-uninfected but who reported one or more symptoms during the study period were also excluded because these symptoms could be indicative of infection by other adventitious agents. SARS-CoV-2-infected participants were considered to be asymptomatic if they did not report any symptoms in the week prior to enrollment and up to 28 days after study enrollment. SARS-CoV-2-infected participants were classified as symptomatic if they reported having fever, cough, or shortness of breath for two or more days; infected participants who reported one or more symptoms during the study period but who did not meet these criteria were excluded from analyses.

### RNA sequencing and bioinformatic processing of sequencing reads

Total RNA was extracted from nasopharyngeal samples using RNeasy kits (Qiagen) and from whole blood using PAXgene blood miRNA extraction kits (Qiagen). RNA quantity and quality were assessed using a NanoDrop 2000 spectrophotometer (Thermo Fisher Scientific, Waltham, MA) and a Bioanalyzer 2100 system with RNA 6000 Nano kits (Agilent, Santa Clara, CA). Median (IQR) RNA integrity numbers for nasopharyngeal and peripheral blood samples were 6.1 (4.7-7.2) and 8.1 (7.5-8.4), respectively. Library preparation was performed using Universal Plus mRNA library kits (Tecan, Männedorf, Switzerland) with AnyDeplete Globin (NuGEN Technologies, Redwood City, CA). The resulting libraries were sequenced on a NovaSeq 6000 instrument (Illumina, San Diego, CA) configured for 100 base-pair paired-end sequencing. Raw sequencing reads were assessed for quality using FastQC v0.11.9 and trimmed using Trimmomatic v0.39 (*53, 54*). Median (IQR) sequencing depth for nasopharyngeal and peripheral blood samples after quality filtering was 41.6 (33.8-55.4) and 49.0 (39.0-55.7) million reads, respectively. The resulting quality-filtered reads were aligned to the human reference genome GRCh38 and a gene count matrix was generated using STAR v2.7.10b (*55*). Gene annotation was performed using the GENCODE v43 basic annotation (*56*). Genes not expressed at a level of at least 1 count per million in at least 50% of samples were excluded from further analyses.

### Statistical analyses

We used Chi-squared or Fisher’s exact tests (categorical variables) and Wilcoxon rank-sum or Kruskal-Wallis tests (continuous variables) to evaluate for differences in sociodemographic and clinical characteristics by age group and SARS-CoV-2 infection status. To estimate the proportions of immune cell populations in upper respiratory and peripheral blood samples, we used the CIBERSORT deconvolution algorithm and the LM22 gene signature matrix (*29*). Briefly, CIBERSORT is an analytical tool that uses support vector regression to estimate the relative proportions of cell types using gene expression data. For this analysis, CIBERSORT was run in relative mode with B-mode batch correction and the number of permutations set to 500. LM22 is a validated gene signature matrix consisting of 547 genes that have been shown to accurately differentiate 22 immune cell populations, including 11 major leukocyte types (*29*). We compared the proportions of immune cell populations by age (modeled as a continuous variable) and SARS-CoV-2 infection status using beta regression (*57*). We observed substantial differences in immune cell populations in both upper respiratory and peripheral blood samples by these variables; thus, we chose to adjust all gene expression analyses for sample immune cell composition. To reduce the dimensionality of these data, we generated principal components separately for upper respiratory and peripheral blood samples using data for the 11 major leukocyte types in the LM22 signature matrix. We then adjusted gene expression analyses for the first five principal components generated for each sample type, accounting for 77% and 83% of the variance in the proportions of major leukocyte types across upper respiratory and peripheral blood samples, respectively. We additionally adjusted these analyses for sequencing batch and participant sex and, when not the comparison of interest, we adjusted for or stratified by age group and SARS-CoV-2 infection status. We used DESeq2 to evaluate for differences in the expression of individual genes across groups of interest (*58*). We identified modules comprised of groups of co-expressed genes that share a similar function using the gene sets defined by the NanoString nCounter^®^ Host Response Panel (lists of genes for each module are provided in the **Supplemental Materials**) (*31*). We then used fast gene set enrichment analysis (FGSEA) to evaluate for differential expression of these modules across comparisons of interest (*30*). Finally, we used single-sample gene set enrichment analysis (ssGSEA) to generate module enrichment scores for each sample among SARS-CoV-2-infected children and adolescents with paired upper respiratory and peripheral blood samples. We then calculated Pearson’s correlation coefficients to evaluate for linear relationships between expression of these modules in the URT and peripheral blood of the same individual (*59*). Corrections for the false discovery rate due to multiple testing were performed using the Benjamini-Hochberg procedure. All analyses were performed using R version 4.3.1 (*60*).

#### Study approval

The BRAVE Kids study is being conducted within the Duke University Health System (DUHS) in Raleigh-Durham, North Carolina, USA. DUHS is a large, integrated health system consisting of 3 hospitals and over 100 outpatient clinics. This study was approved by the DUHS Institutional Review Board (Pro00106150). Informed consent was obtained from all study participants or their legal guardians, with written approval obtained using an electronic consent document. The MESSI study was approved by the DUHS Institutional Review Board (Pro00100241). Informed consent was obtained from all study participants, with written approval obtained using an electronic consent document.

## Supporting information

nCounter Host Response Gene List

## Data Availability

All data are available in the main text or the supplementary materials. The sequencing dataset supporting the conclusions of this study is available in the Gene Expression Omnibus (accession number: GSE231409). The statistical files and script used for data analyses are also publicly available (https://github.com/mskelly7/COVID_RNASeq_Age).

## List of Supplementary Materials

nCounter Host Response Gene List

Fig. S1 to S5

Tables S1 to S8

## Acknowledgments

We offer sincere gratitude to the children and families who participated in this research.

## Author contributions

Conceptualization: JHH, MTM, RH, CWW, MSK

Methodology: RH, KMW, MSK

Investigation: JHH, AAM, TD, IG, JNA, DJL, TSP, JR, ATR, NAT, TWB, MTM, RH, CTD, RL, TND, KMW, ZX, AM, OR, CWW, MSK

Visualization: AAM, MSK

Funding acquisition: MSK

Project administration: JHH, CWW, MSK

Supervision: MSK

Writing – original draft: JHH, MSK

Writing – review & editing: JHH, AAM, TD, IG, JNA, DJL, TSP, JR, ATR, NAT, TWB, MTM, RH, CTD, RL, TND, KMW, ZX, AM, OR, CWW, MSK

## REFERENCES AND NOTES

1. R. Verity, L. C. Okell, I. Dorigatti, P. Winskill, C. Whittaker, N. Imai, G. Cuomo-Dannenburg, H. Thompson, P. G. T. Walker, H. Fu, A. Dighe, J. T. Griffin, M. Baguelin, S. Bhatia, A. Boonyasiri, A. Cori, Z. Cucunubá, R. FitzJohn, K. Gaythorpe, W. Green, A. Hamlet, W. Hinsley, D. Laydon, G. Nedjati-Gilani, S. Riley, S. van Elsland, E. Volz, H. Wang, Y. Wang, X. Xi, C. A. Donnelly, A. C. Ghani, N. M. Ferguson, Estimates of the severity of coronavirus disease 2019: a model-based analysis. Lancet Infect Dis 20, 669–677 (2020).

2. Centers for Disease Control and Prevention (CDC), United States COVID-19 Cases and Deaths by State over Time (available at https://data.cdc.gov/Case-Surveillance/United-States-COVID-19-Cases-and-Deaths-by-State-o/9mfq-cb36).

3. CDC, Cases, Data, and SurveillanceCenters for Disease Control and Prevention (2020) (available at https://www.cdc.gov/coronavirus/2019-ncov/covid-data/investigations-discovery/hospitalization-death-by-age.html).

4. Centers for Disease Control and Prevention, COVID-NET: COVID-19-Associated Hospitalization Surveillance Network (available at https://gis.cdc.gov/grasp/covidnet/COVID19_3.html).

5. E. Molteni, C. H. Sudre, L. S. Canas, S. S. Bhopal, R. C. Hughes, M. Antonelli, B. Murray, K. Kläser, E. Kerfoot, L. Chen, J. Deng, C. Hu, S. Selvachandran, K. Read, J. Capdevila Pujol, A. Hammers, T. D. Spector, S. Ourselin, C. J. Steves, M. Modat, M. Absoud, E. L. Duncan, Illness duration and symptom profile in symptomatic UK school-aged children tested for SARS-CoV-2. The Lancet Child & Adolescent Health 5, 708–718 (2021).

6. J. W. Antoon, C. G. Grijalva, C. Thurm, T. Richardson, A. B. Spaulding, R. J. T. Ii, M. A. Reyes, S. S. Shah, J. E. Burns, C. C. Kenyon, A. L. Hersh, D. J. Williams, Factors Associated With COVIDD19 Disease Severity in US Children and Adolescents. Journal of Hospital Medicine 16, 603–610 (2021).

7. J. H. Hurst, S. M. Heston, H. N. Chambers, H. M. Cunningham, M. J. Price, L. Suarez, C. G. Crew, S. Bose, J. N. Aquino, S. T. Carr, S. M. Griffin, S. H. Smith, K. Jenkins, T. S. Pfeiffer, J. Rodriguez, C. T. DeMarco, N. A. De Naeyer, T. C. Gurley, R. Louzao, C. Zhao, C. K. Cunningham, W. J. Steinbach, T. N. Denny, D. J. Lugo, M. A. Moody, S. R. Permar, A. T. Rotta, N. A. Turner, E. B. Walter, C. W. Woods, M. S. Kelly, Severe Acute Respiratory Syndrome Coronavirus 2 Infections Among Children in the Biospecimens from Respiratory Virus-Exposed Kids (BRAVE Kids) Study. Clin Infect Dis 73, e2875–e2882 (2021).

8. M. K. Steele, A. Couture, C. Reed, D. Iuliano, M. Whitaker, H. Fast, A. J. Hall, A. MacNeil, B. Cadwell, K. J. Marks, B. J. Silk, Estimated Number of COVID-19 Infections, Hospitalizations, and Deaths Prevented Among Vaccinated Persons in the US, December 2020 to September 2021. JAMA Netw Open 5, e2220385 (2022).

9. CDC, COVID Data TrackerCenters for Disease Control and Prevention (2020) (available at https://covid.cdc.gov/covid-data-tracker).

10. S. Adjei, K. Hong, N.-A. M. Molinari, L. Bull-Otterson, U. A. Ajani, A. V. Gundlapalli, A. M. Harris, J. Hsu, S. S. Kadri, J. Starnes, K. Yeoman, T. K. Boehmer, Mortality Risk Among Patients Hospitalized Primarily for COVID-19 During the Omicron and Delta Variant Pandemic Periods — United States, April 2020–June 2022. MMWR Morb. Mortal. Wkly. Rep. 71, 1182–1189 (2022).

11. F. Zhou, T. Yu, R. Du, G. Fan, Y. Liu, Z. Liu, J. Xiang, Y. Wang, B. Song, X. Gu, L. Guan, Y. Wei, H. Li, X. Wu, J. Xu, S. Tu, Y. Zhang, H. Chen, B. Cao, Clinical course and risk factors for mortality of adult inpatients with COVID-19 in Wuhan, China: a retrospective cohort study. Lancet 395, 1054–1062 (2020).

12. M. Miyara, M. Saichi, D. Sterlin, F. Anna, S. Marot, A. Mathian, M. Atif, P. Quentric, A. Mohr, L. Claër, C. Parizot, K. Dorgham, H. Yssel, J. Fadlallah, T. Chazal, J. Haroche, C.-E. Luyt, J. Mayaux, A. Beurton, N. Benameur, D. Boutolleau, S. Burrel, S. de Alba, S. Mudumba, R. Hockett, C. Gunn, P. Charneau, V. Calvez, A.-G. Marcelin, A. Combes, A. Demoule, Z. Amoura, G. Gorochov, Pre-COVID-19 Immunity to Common Cold Human Coronaviruses Induces a Recall-Type IgG Response to SARS-CoV-2 Antigens Without Cross-Neutralisation. Front Immunol 13, 790334 (2022).

13. S. Bunyavanich, A. Do, A. Vicencio, Nasal Gene Expression of Angiotensin-Converting Enzyme 2 in Children and Adults. JAMA 323, 2427–2429 (2020).

14. Z. Zhang, L. Guo, L. Huang, C. Zhang, R. Luo, L. Zeng, H. Liang, Q. Li, X. Lu, X. Wang, C. Y. Ma, J. Shao, W. Luo, L. Li, L. Liu, Z. Li, X. Zhou, X. Zhang, J. Liu, J. Yang, K. Y. Kwan, W. Liu, Y. Xu, H. Jiang, H. Liu, H. Du, Y. Wu, G. Yu, J. Chen, J. Wu, J. Zhang, C. Liao, H. J. Chen, Z. Chen, H. Tse, H. Xia, Q. Lian, Distinct Disease Severity Between Children and Older Adults With Coronavirus Disease 2019 (COVID-19): Impacts of ACE2 Expression, Distribution, and Lung Progenitor Cells. Clinical Infectious Diseases 73, e4154–e4165 (2021).

15. S. Kadambari, P. Klenerman, A. J. Pollard, Why the elderly appear to be more severely affected by COVID-19: The potential role of immunosenescence and CMV. Rev Med Virol 30, e2144 (2020).

16. V. Bajaj, N. Gadi, A. P. Spihlman, S. C. Wu, C. H. Choi, V. R. Moulton, Aging, Immunity, and COVID-19: How Age Influences the Host Immune Response to Coronavirus Infections? Frontiers in Physiology 11 (2021) (available at https://www.frontiersin.org/articles/10.3389/fphys.2020.571416).

17. J. W. Prokop, N. L. Hartog, D. Chesla, W. Faber, C. P. Love, R. Karam, N. Abualkheir, B. Feldmann, L. Teng, T. McBride, M. L. Leimanis, B. K. English, A. Holsworth, A. Frisch, J. Bauss, N. Kalpage, A. Derbedrossian, R. M. Pinti, N. Hale, J. Mills, A. Eby, E. A. VanSickle, S. C. Pageau, R. Shankar, B. Chen, J. A. Carcillo, D. Sanfilippo, R. Olivero, C. P. Bupp, S. Rajasekaran, High-Density Blood Transcriptomics Reveals Precision Immune Signatures of SARS-CoV-2 Infection in Hospitalized Individuals. Front. Immunol. 12, 694243 (2021).

18. P. Bost, F. De Sanctis, S. Canè, S. Ugel, K. Donadello, M. Castellucci, D. Eyal, A. Fiore, C. Anselmi, R. M. Barouni, R. Trovato, S. Caligola, A. Lamolinara, M. Iezzi, F. Facciotti, A. Mazzariol, D. Gibellini, P. De Nardo, E. Tacconelli, L. Gottin, E. Polati, B. Schwikowski, I. Amit, V. Bronte, Deciphering the state of immune silence in fatal COVID-19 patients. Nat Commun 12, 1428 (2021).

19. Y. Hou, Y. Zhou, L. Jehi, Y. Luo, M. U. Gack, T. A. Chan, H. Yu, C. Eng, A. A. Pieper, F. Cheng, AgingDrelated cell typeDspecific pathophysiologic immune responses that exacerbate disease severity in aged COVIDD19 patients. Aging Cell 21 (2022), doi:10.1111/acel.13544.

20. M. T. McClain, F. J. Constantine, R. Henao, Y. Liu, E. L. Tsalik, T. W. Burke, J. M. Steinbrink, E. Petzold, B. P. Nicholson, R. Rolfe, B. D. Kraft, M. S. Kelly, D. R. Saban, C. Yu, X. Shen, E. M. Ko, G. D. Sempowski, T. N. Denny, G. S. Ginsburg, C. W. Woods, Dysregulated transcriptional responses to SARS-CoV-2 in the periphery. Nat Commun 12, 1079 (2021).

21. E. A. Oliveira, E. A. Colosimo, A. C. Simões e Silva, R. H. Mak, D. B. Martelli, L. R. Silva, H. Martelli-Júnior, M. C. L. Oliveira, Clinical characteristics and risk factors for death among hospitalised children and adolescents with COVID-19 in Brazil: an analysis of a nationwide database. The Lancet Child & Adolescent Health 5, 559–568 (2021).

22. S. Bellino, O. Punzo, M. C. Rota, M. Del Manso, A. M. Urdiales, X. Andrianou, M. Fabiani, S. Boros, F. Vescio, F. Riccardo, A. Bella, A. Filia, G. Rezza, A. Villani, P. Pezzotti, COVID-19 WORKING GROUP, COVID-19 Disease Severity Risk Factors for Pediatric Patients in Italy. Pediatrics 146, e2020009399 (2020).

23. M. Ilieva, M. Tschaikowski, A. Vandin, S. Uchida, The current status of gene expression profilings in COVIDD19 patients. Clinical and Translational Dis 2 (2022), doi:10.1002/ctd2.104.

24. M. Yoshida, K. B. Worlock, N. Huang, R. G. H. Lindeboom, C. R. Butler, N. Kumasaka, C. Dominguez Conde, L. Mamanova, L. Bolt, L. Richardson, K. Polanski, E. Madissoon, J. L. Barnes, J. Allen-Hyttinen, E. Kilich, B. C. Jones, A. de Wilton, A. Wilbrey-Clark, W. Sungnak, J. P. Pett, J. Weller, E. Prigmore, H. Yung, P. Mehta, A. Saleh, A. Saigal, V. Chu, J. M. Cohen, C. Cane, A. Iordanidou, S. Shibuya, A.-K. Reuschl, I. T. Herczeg, A. C. Argento, R. G. Wunderink, S. B. Smith, T. A. Poor, C. A. Gao, J. E. Dematte, NU SCRIPT Study Investigators, G. R. S. Budinger, H. K. Donnelly, N. S. Markov, Z. Lu, G. Reynolds, M. Haniffa, G. S. Bowyer, M. Coates, M. R. Clatworthy, F. J. Calero-Nieto, B. Göttgens, C. O’Callaghan, N. J. Sebire, C. Jolly, P. De Coppi, C. M. Smith, A. V. Misharin, S. M. Janes, S. A. Teichmann, M. Z. Nikolić, K. B. Meyer, Local and systemic responses to SARS-CoV-2 infection in children and adults. Nature 602, 321–327 (2022).

25. C. A. Pierce, S. Sy, B. Galen, D. Y. Goldstein, E. Orner, M. J. Keller, K. C. Herold, B. C. Herold, Natural mucosal barriers and COVID-19 in children. JCI Insight 6, e148694 (2021).

26. C. M. Koch, A. D. Prigge, K. R. Anekalla, A. Shukla, H. C. Do Umehara, L. Setar, J. Chavez, H. Abdala-Valencia, Y. Politanska, N. S. Markov, G. R. Hahn, T. Heald-Sargent, L. N. Sanchez-Pinto, W. J. Muller, B. D. Singer, A. V. Misharin, K. M. Ridge, B. M. Coates, Age-related Differences in the Nasal Mucosal Immune Response to SARS-CoV-2. Am J Respir Cell Mol Biol 66, 206–222 (2022).

27. A. Georgountzou, N. G. Papadopoulos, Postnatal Innate Immune Development: From Birth to Adulthood. Frontiers in Immunology 8 (2017) (available at https://www.frontiersin.org/articles/10.3389/fimmu.2017.00957).

28. S. Jalali, C. M. Harpur, A. T. Piers, M. Auladell, L. Perriman, S. Li, K. An, J. Anderson, S. P. Berzins, P. V. Licciardi, T. M. Ashhurst, I. E. Konstantinov, D. G. Pellicci, A high-dimensional cytometry atlas of peripheral blood over the human life span. Immunology & Cell Biology 100, 805–821 (2022).

29. A. M. Newman, C. B. Steen, C. L. Liu, A. J. Gentles, A. A. Chaudhuri, F. Scherer, M. S. Khodadoust, M. S. Esfahani, B. A. Luca, D. Steiner, M. Diehn, A. A. Alizadeh, Determining cell type abundance and expression from bulk tissues with digital cytometry. Nat Biotechnol 37, 773–782 (2019).

30. G. Korotkevich, V. Sukhov, N. Budin, B. Shpak, M. N. Artyomov, A. Sergushichev, Fast gene set enrichment analysis (Bioinformatics, 2016; http://biorxiv.org/lookup/doi/10.1101/060012).

31. Host ResponseNanoString (2023) (available at https://nanostring.com/products/ncounter-assays-panels/immunology/host-response/).

32. C. Rydyznski Moderbacher, S. I. Ramirez, J. M. Dan, A. Grifoni, K. M. Hastie, D. Weiskopf, S. Belanger, R. K. Abbott, C. Kim, J. Choi, Y. Kato, E. G. Crotty, C. Kim, S. A. Rawlings, J. Mateus, L. P. V. Tse, A. Frazier, R. Baric, B. Peters, J. Greenbaum, E. Ollmann Saphire, D. M. Smith, A. Sette, S. Crotty, Antigen-Specific Adaptive Immunity to SARS-CoV-2 in Acute COVID-19 and Associations with Age and Disease Severity. Cell 183, 996–1012.e19 (2020).

33. S. Felsenstein, C. M. Hedrich, SARS-CoV-2 infections in children and young people. Clinical Immunology 220, 108588 (2020).

34. D. Herrera-Esposito, G. de los Campos, Age-specific rate of severe and critical SARS-CoV-2 infections estimated with multi-country seroprevalence studies. BMC Infect Dis 22, 311 (2022).

35. M. Vono, A. Huttner, S. Lemeille, P. Martinez-Murillo, B. Meyer, S. Baggio, S. Sharma, A. Thiriard, A. Marchant, G.-J. Godeke, C. Reusken, C. Alvarez, F. Perez-Rodriguez, I. Eckerle, L. Kaiser, N. Loevy, C. S. Eberhardt, G. Blanchard-Rohner, C.-A. Siegrist, A. M. Didierlaurent, Robust innate responses to SARS-CoV-2 in children resolve faster than in adults without compromising adaptive immunity. Cell Reports 37, 109773 (2021).

36. C. A. Pierce, P. Preston-Hurlburt, Y. Dai, C. B. Aschner, N. Cheshenko, B. Galen, S. J. Garforth, N. G. Herrera, R. K. Jangra, N. C. Morano, E. Orner, S. Sy, K. Chandran, J. Dziura, S. C. Almo, A. Ring, M. J. Keller, K. C. Herold, B. C. Herold, Immune responses to SARS-CoV-2 infection in hospitalized pediatric and adult patients. Sci. Transl. Med. 12, eabd5487 (2020).

37. R. Gu, T. Mao, Q. Lu, T. Tianjiao Su, J. Wang, Myeloid dysregulation and therapeutic intervention in COVID-19. Semin Immunol 55, 101524 (2021).

38. J. Schulte-Schrepping, N. Reusch, D. Paclik, K. Baßler, S. Schlickeiser, B. Zhang, B. Krämer, T. Krammer, S. Brumhard, L. Bonaguro, E. De Domenico, D. Wendisch, M. Grasshoff, T. S. Kapellos, M. Beckstette, T. Pecht, A. Saglam, O. Dietrich, H. E. Mei, A. R. Schulz, C. Conrad, D. Kunkel, E. Vafadarnejad, C.-J. Xu, A. Horne, M. Herbert, A. Drews, C. Thibeault, M. Pfeiffer, S. Hippenstiel, A. Hocke, H. Müller-Redetzky, K.-M. Heim, F. Machleidt, A. Uhrig, L. Bosquillon de Jarcy, L. Jürgens, M. Stegemann, C. R. Glösenkamp, H.-D. Volk, C. Goffinet, M. Landthaler, E. Wyler, P. Georg, M. Schneider, C. Dang-Heine, N. Neuwinger, K. Kappert, R. Tauber, V. Corman, J. Raabe, K. M. Kaiser, M. T. Vinh, G. Rieke, C. Meisel, T. Ulas, M. Becker, R. Geffers, M. Witzenrath, C. Drosten, N. Suttorp, C. von Kalle, F. Kurth, K. Händler, J. L. Schultze, A. C. Aschenbrenner, Y. Li, J. Nattermann, B. Sawitzki, A.-E. Saliba, L. E. Sander, Severe COVID-19 Is Marked by a Dysregulated Myeloid Cell Compartment. Cell 182, 1419–1440.e23 (2020).

39. S. T. Chen, M. D. Park, D. M. Del Valle, M. Buckup, A. Tabachnikova, R. C. Thompson, N. W. Simons, K. Mouskas, B. Lee, D. Geanon, D. D’Souza, T. Dawson, R. Marvin, K. Nie, Z. Zhao, J. LeBerichel, C. Chang, H. Jamal, G. Akturk, U. Chaddha, K. Mathews, S. Acquah, S.-A. Brown, M. Reiss, T. Harkin, M. Feldmann, C. A. Powell, J. L. Hook, S. Kim-Schulze, A. H. Rahman, B. D. Brown, Mount Sinai COVID-19 Biobank Team, N. D. Beckmann, S. Gnjatic, E. Kenigsberg, A. W. Charney, M. Merad, A shift in lung macrophage composition is associated with COVID-19 severity and recovery. Sci Transl Med 14, eabn5168 (2022).

40. P. Zimmermann, N. Curtis, Why is COVID-19 less severe in children? A review of the proposed mechanisms underlying the age-related difference in severity of SARS-CoV-2 infections. Arch Dis Child (2020), doi:10.1136/archdischild-2020-320338.

41. A. Iwasaki, E. F. Foxman, R. D. Molony, Early local immune defenses in the respiratory tract. Nat Rev Immunol 17, 7–20 (2017).

42. J. Loske, J. Röhmel, S. Lukassen, S. Stricker, V. G. Magalhães, J. Liebig, R. L. Chua, L. Thürmann, M. Messingschlager, A. Seegebarth, B. Timmermann, S. Klages, M. Ralser, B. Sawitzki, L. E. Sander, V. M. Corman, C. Conrad, S. Laudi, M. Binder, S. Trump, R. Eils, M. A. Mall, I. Lehmann, Pre-activated antiviral innate immunity in the upper airways controls early SARS-CoV-2 infection in children. Nat Biotechnol 40, 319–324 (2022).

43. K. Oishi, S. Horiuchi, J. Frere, R. E. Schwartz, B. R. tenOever, A diminished immune response underlies age-related SARS-CoV-2 pathologies. Cell Reports 39, 111002 (2022).

44. Y.-I. Kim, K.-M. Yu, J.-Y. Koh, E.-H. Kim, S.-M. Kim, E. J. Kim, M. A. B. Casel, R. Rollon, S.-G. Jang, M.-S. Song, S.-J. Park, H. W. Jeong, E.-G. Kim, O.-J. Lee, Y.-D. Kim, Y. Choi, S.-A. Lee, Y. J. Choi, S.-H. Park, J. U. Jung, Y. K. Choi, Age-dependent pathogenic characteristics of SARS-CoV-2 infection in ferrets. Nat Commun 13, 21 (2022).

45. N. R. Cheemarla, T. A. Watkins, V. T. Mihaylova, B. Wang, D. Zhao, G. Wang, M. L. Landry, E. F. Foxman, Dynamic innate immune response determines susceptibility to SARS-CoV-2 infection and early replication kinetics. Journal of Experimental Medicine 218, e20210583 (2021).

46. X. An, M. Martinez-Paniagua, A. Rezvan, S. R. Sefat, M. Fathi, S. Singh, S. Biswas, M. Pourpak, C. Yee, X. Liu, N. Varadarajan, Single-dose intranasal vaccination elicits systemic and mucosal immunity against SARS-CoV-2. iScience 24, 103037 (2021).

47. A. O. Hassan, F. Feldmann, H. Zhao, D. T. Curiel, A. Okumura, T.-L. Tang-Huau, J. B. Case, K. Meade-White, J. Callison, R. E. Chen, J. Lovaglio, P. W. Hanley, D. P. Scott, D. H. Fremont, H. Feldmann, M. S. Diamond, A single intranasal dose of chimpanzee adenovirus-vectored vaccine protects against SARS-CoV-2 infection in rhesus macaques. Cell Reports Medicine 2, 100230 (2021).

48. M. A. Wolf, D. T. Boehm, M. A. DeJong, T. Y. Wong, E. Sen-Kilic, J. M. Hall, C. B. Blackwood, K. L. Weaver, C. O. Kelly, C. A. Kisamore, G. J. Bitzer, J. R. Bevere, M. Barbier, F. H. Damron, Intranasal Immunization with Acellular Pertussis Vaccines Results in Long-Term Immunity to Bordetella pertussis in Mice. Infection and Immunity 89, 10.1128/iai.00607-20 (2021).

49. C. M. Oshansky, A. J. Gartland, S.-S. Wong, T. Jeevan, D. Wang, P. L. Roddam, M. A. Caniza, T. Hertz, J. P. DeVincenzo, R. J. Webby, P. G. Thomas, Mucosal Immune Responses Predict Clinical Outcomes during Influenza Infection Independently of Age and Viral Load. Am J Respir Crit Care Med 189, 449–462 (2014).

50. C. García, A. Soriano-Fallas, J. Lozano, N. Leos, A. M. Gomez, O. Ramilo, A. Mejias, Decreased innate immune cytokine responses correlate with disease severity in children with respiratory syncytial virus and human rhinovirus bronchiolitis. Pediatr Infect Dis J 31, 86–89 (2012).

51. J. Taveras, C. Garcia-Maurino, M. Moore-Clingenpeel, Z. Xu, S. Mertz, F. Ye, P. Chen, S. H. Cohen, D. Cohen, M. E. Peeples, O. Ramilo, A. Mejias, Type III Interferons, Viral Loads, Age, and Disease Severity in Young Children With Respiratory Syncytial Virus Infection. J Infect Dis 227, 61–70 (2022).

52. N. Smith, P. Goncalves, B. Charbit, L. Grzelak, M. Beretta, C. Planchais, T. Bruel, V. Rouilly, V. Bondet, J. Hadjadj, N. Yatim, H. Pere, S. H. Merkling, A. Ghozlane, S. Kernéis, F. Rieux-Laucat, B. Terrier, O. Schwartz, H. Mouquet, D. Duffy, J. P. Di Santo, Distinct systemic and mucosal immune responses during acute SARS-CoV-2 infection. Nat Immunol 22, 1428–1439 (2021).

53. Babraham Bioinformatics - FastQC A Quality Control tool for High Throughput Sequence Data (available at https://www.bioinformatics.babraham.ac.uk/projects/fastqc/).

54. A. M. Bolger, M. Lohse, B. Usadel, Trimmomatic: a flexible trimmer for Illumina sequence data. Bioinformatics 30, 2114–2120 (2014).

55. A. Dobin, C. A. Davis, F. Schlesinger, J. Drenkow, C. Zaleski, S. Jha, P. Batut, M. Chaisson, T. R. Gingeras, STAR: ultrafast universal RNA-seq aligner. Bioinformatics 29, 15–21 (2013).

56. A. Frankish, M. Diekhans, A.-M. Ferreira, R. Johnson, I. Jungreis, J. Loveland, J. M. Mudge, C. Sisu, J. Wright, J. Armstrong, I. Barnes, A. Berry, A. Bignell, S. Carbonell Sala, J. Chrast, F. Cunningham, T. Di Domenico, S. Donaldson, I. T. Fiddes, C. García Girón, J. M. Gonzalez, T. Grego, M. Hardy, T. Hourlier, T. Hunt, O. G. Izuogu, J. Lagarde, F. J. Martin, L. Martínez, S. Mohanan, P. Muir, F. C. P. Navarro, A. Parker, B. Pei, F. Pozo, M. Ruffier, B. M. Schmitt, E. Stapleton, M.-M. Suner, I. Sycheva, B. Uszczynska-Ratajczak, J. Xu, A. Yates, D. Zerbino, Y. Zhang, B. Aken, J. S. Choudhary, M. Gerstein, R. Guigó, T. J. P. Hubbard, M. Kellis, B. Paten, A. Reymond, M. L. Tress, P. Flicek, GENCODE reference annotation for the human and mouse genomes. Nucleic Acids Research 47, D766–D773 (2019).

57. J. C. Douma, J. T. Weedon, Analysing continuous proportions in ecology and evolution: A practical introduction to beta and Dirichlet regression. Methods in Ecology and Evolution 10, 1412–1430 (2019).

58. M. Love, W. Huber, S. Anders, Moderated estimation of fold change and dispersion for RNA-seq data with DESeq2. Genome Biology 15, 550 (2014).

59. X. Rui, S. Shao, L. Wang, J. Leng, Identification of recurrence marker associated with immune infiltration in prostate cancer with radical resection and build prognostic nomogram. BMC Cancer 19, 1179 (2019).

60. R: The R Project for Statistical Computing (available at https://www.r-project.org/).

