## Supplemental Figures for "Differential host responses within the upper respiratory tract and peripheral blood of children and adults with SARS-CoV-2 infection"

**Supplemental Materials**

**nCounter Host Response Gene List.** Gene modules used for gene set enrichment analysis were based on modules defined by the NanoString nCounter^®^ Host Response Panel. A full list of the genes included in each module is provided.

**Supplemental Figures**

Fig. S1 – Fig. S5

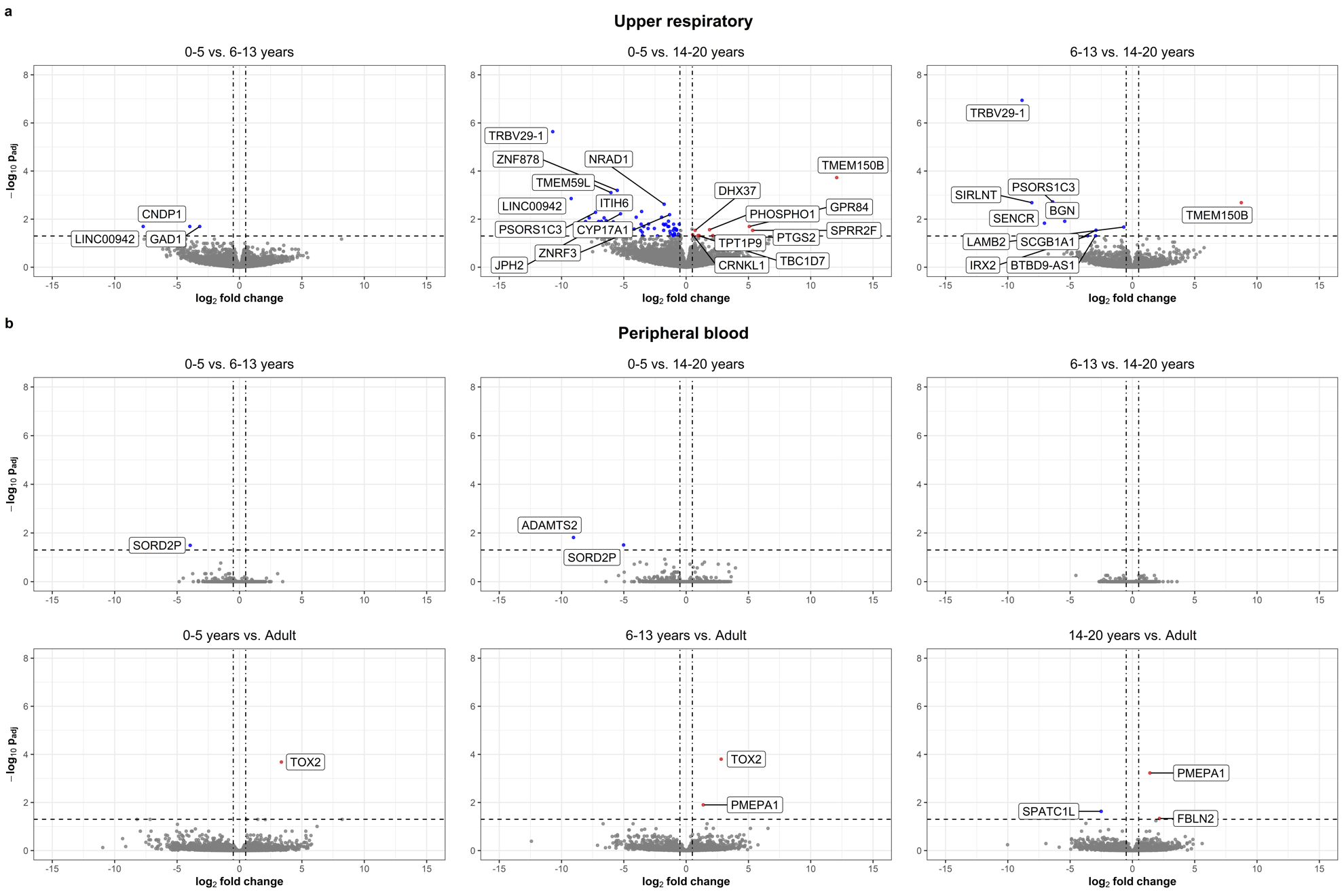

**Fig. S1.** **Differential expression of individual genes among SARS-CoV-2-uninfected pediatric subjects by age group.** Volcano plots are shown comparing the transcriptional profiles of upper respiratory (**a.**) and peripheral blood (**b.**) samples from SARS-CoV-2-uninfected young children (0-5 years), school-age children (6-13 years), adolescents (14-20 years), and adults (≥21 years, peripheral blood only). For each comparison, the age group listed first represents the group of interest while the age group listed second is the reference group. Differentially expressed genes are colored red (upregulated in the age group of interest) or blue (downregulated in the age group of interest). When applicable, the 10 most differentially upregulated and downregulated genes based on log2-fold change are labeled. All analyses were adjusted for sex, sequencing batch, and imputed sample immune cell proportions.

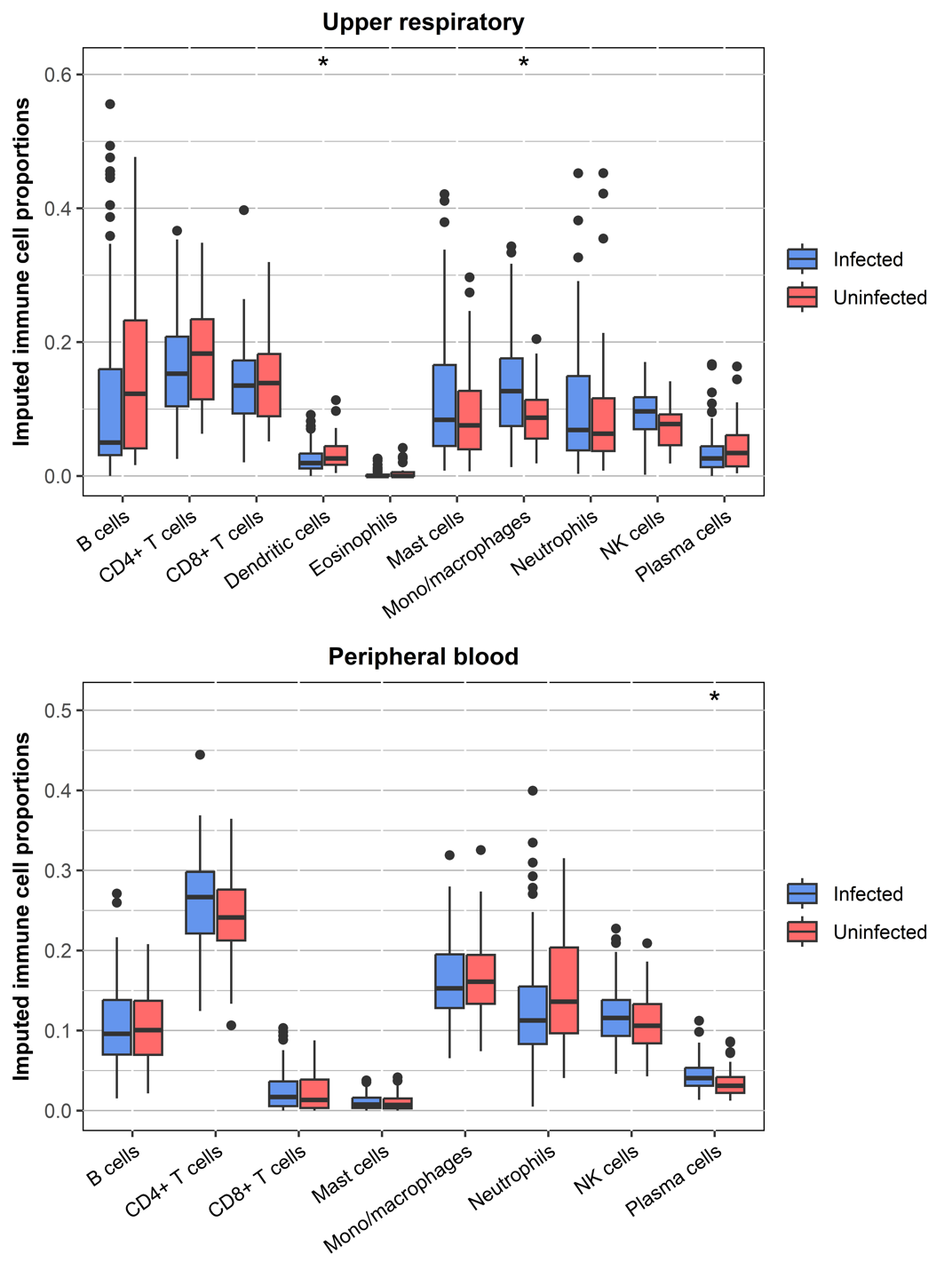

**Fig. S2.** **Imputed sample immune cell proportions in children, adolescents, and adults with SARS-CoV-2 infection and uninfected controls.** Bulk RNA sequencing was used to compare the transcriptional profiles of SARS-CoV-2-infected and uninfected young children (0-5 years), school-age children (6-13 years), adolescents (14-20 years) and adults (≥21 years, peripheral blood only). Box and whisker plots depict proportions of immune cell populations imputed using CIBERSORT in upper respiratory (**a.**) and peripheral blood (**b.**) Lines splitting the boxes correspond to median values while box edges represent the 25^th^ and 75^th^ percentiles with outliers shown as single points. Data for SARS-CoV-2-infected and uninfected subjects are shown in blue and red, respectively. Proportions of immune cell populations were compared by SARS-CoV-2 status using beta regression, with all analyses adjusted for age (modeled as a continuous variable) and corrected for multiple comparisons (*, *p*_adj_<0.05; **, *p*_adj_<0.01; ***, *p*_adj_<0.001; ****, *p*_adj_<0.0001). Only immune cell populations identified in at least 25% of samples are shown.

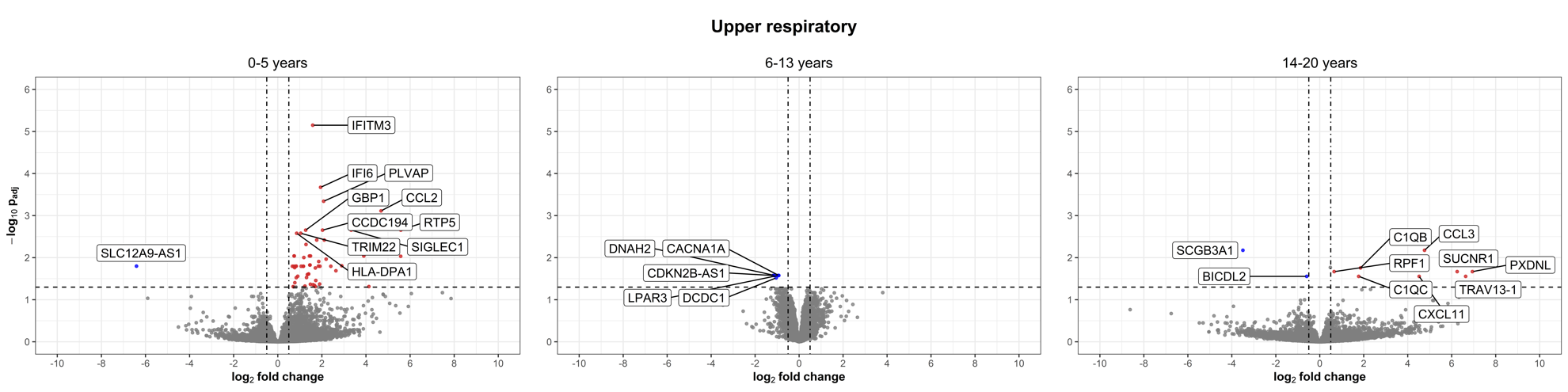

**Fig. S3.** **Differential expression of individual genes in the upper respiratory tract associated with SARS-CoV-2 infection among children and adolescents.** Bulk RNA sequencing was used to compare the upper respiratory transcriptional profiles of young children (0-5 years), school-age children (6-13 years), and adolescents (14-20 years) by SARS-CoV-2 infection status. Volcano plots are shown depicting differential expression of genes among SARS-CoV-2-infected subjects relative to uninfected subjects by age group (*p*_adj_<0.05). When applicable, the 10 most differentially upregulated and downregulated genes based on log2-fold change are labeled. All analyses were adjusted for sex, sequencing batch, and imputed sample immune cell proportions.

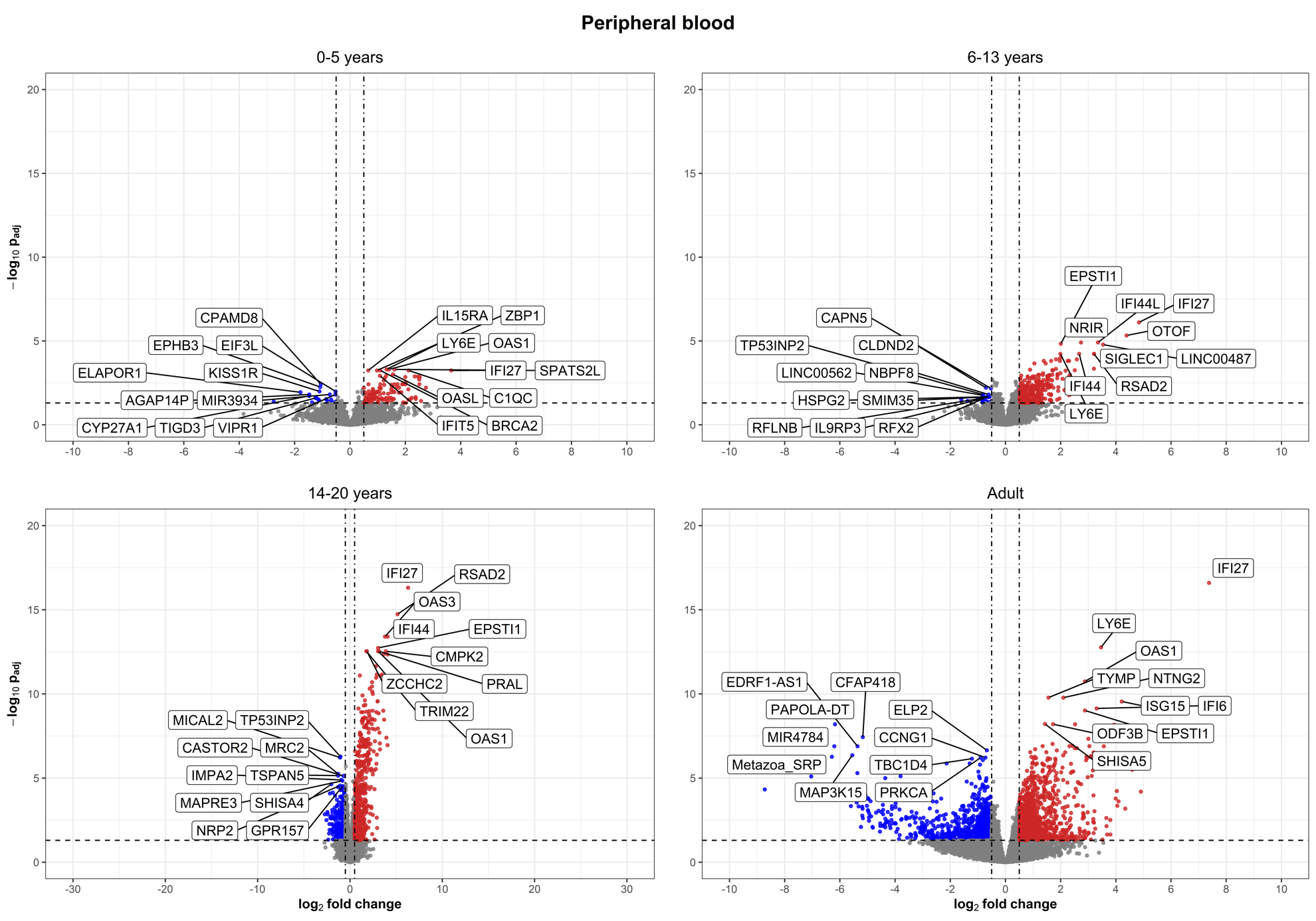

**Fig. S4.** **Differential expression of individual genes in peripheral blood associated with SARS-CoV-2 infection among children, adolescents, and adults.** Bulk RNA sequencing was used to compare the peripheral blood transcriptional profiles of young children (0-5 years), school-age children (6-13 years), adolescents (14-20 years), and adults (≥21 years) by SARS-CoV-2 infection status. Volcano plots are shown depicting differential expression of genes among SARS-CoV-2-infected subjects relative to uninfected subjects in the same age group (*p*_adj_<0.05). The 10 most differentially upregulated and downregulated genes based on log2-fold change are labeled. All analyses were adjusted for sex, sequencing batch, and imputed sample immune cell proportions.

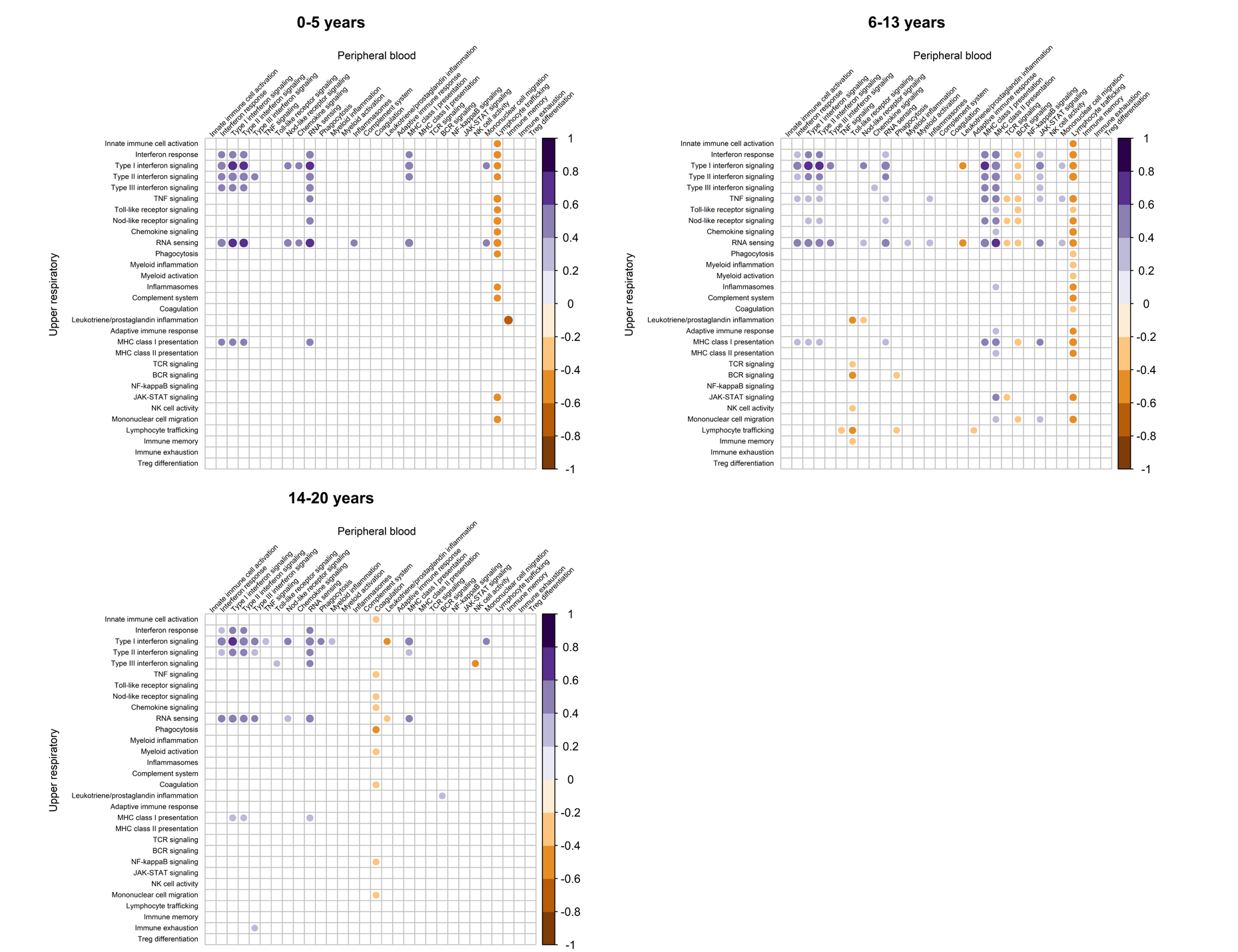

**Fig. S5.** **Correlations between immune module expression in the upper respiratory tract and peripheral blood of SARS-CoV-2-infected children and adolescents by age group.** Single-sample gene set enrichment analysis was used to calculate enrichment scores for gene modules in paired upper respiratory and peripheral blood samples from SARS-CoV-2-infected young children (0-5 years), school-age children (6-13 years), and adolescents (14-20 years). Pearson’s correlation coefficients were then calculated to evaluate for linear relationships between expression of modules in the upper respiratory tract and peripheral blood of the same individual within specific age groups. Positive correlations are displayed in purple and negative correlations are displayed in orange. The size and color of each circle corresponds to the strength of the correlation; only statistically significant correlations (*p*<0.05) are shown.

**Supplemental Tables**

**Table S1.** Output from fast gene set enrichment analysis of immune module expression in upper respiratory tract samples from healthy subjects

**Table S2.** Output from fast gene set enrichment analysis of immune module expression in peripheral blood samples from healthy subjects

**Table S3.** Output from fast gene set enrichment analysis of immune modules in upper respiratory and peripheral blood samples by SARS-CoV-2 infection status

**Table S4.** Output from fast gene set enrichment analysis of immune modules in upper respiratory samples by SARS-CoV-2 infection status within specific age groups

**Table S5.** Output from fast gene set enrichment analysis of immune modules in peripheral blood samples by SARS-CoV-2 infection status within specific age groups

**Table S6.** Output from fast gene set enrichment analysis of immune modules in upper respiratory samples from SARS-CoV-2-infected participants by subject and illness characteristics

**Table S7.** Output from fast gene set enrichment analysis of immune modules in peripheral blood samples from SARS-CoV-2-infected participants by subject and illness characteristics

**Table S8.** Correlations between expression of immune modules in paired upper respiratory and peripheral blood samples collected from the same individual within pediatric age groups.
